# Lexical Markers of Disordered Speech in Primary Progressive Aphasia and ‘Parkinson-plus’ Disorders

**DOI:** 10.1101/2023.11.05.23298112

**Authors:** Shalom K. Henderson, Siddharth Ramanan, Karalyn E. Patterson, Peter Garrard, Nikil Patel, Katie A. Peterson, Ajay Halai, Stefano F. Cappa, James B. Rowe, Matthew A. Lambon Ralph

**Author notes:** Joint senior authors (James B. Rowe, Matthew A. Lambon Ralph). Correspondence to: Shalom K. Henderson, MRC Cognition and Brain Sciences Unit, University of Cambridge, Cambridge CB2 7EF UK. For the purpose of open access, the author has applied a CC BY public copyright licence to any Author Accepted Manuscript version arising from this submission.

## Abstract

Connected speech samples elicited by a picture description task are widely used in the assessment of aphasias, but it is not clear what their interpretation should focus on. Although such samples are easy to collect, analyses of them tend to be time-consuming, inconsistently conducted, and impractical for non-specialist settings. Here, we analysed connected speech samples from patients with the three variants of primary progressive aphasia (semantic, svPPA N = 9; logopenic, lvPPA N = 9; non-fluent, nfvPPA N = 9), progressive supranuclear palsy (PSP Richardson’s syndrome N = 10), corticobasal syndrome (CBS N = 13), and age-matched healthy controls. There were three principal aims: (i) to determine the differences in quantitative language output and psycholinguistic properties of words produced by patients and controls; (ii) to identify the neural correlates of connected speech measures; and (iii) to develop a simple clinical measurement tool. Using data-driven methods, we optimised a 15-word checklist for use with the Boston Diagnostic Aphasia Examination ‘cookie theft’ and Mini Linguistic State Examination ‘beach scene’ pictures and tested the predictive validity of outputs from *Least Absolute Shrinkage and Selection Operator* (LASSO) models using an independent clinical sample from a second site. The total language output was significantly reduced in patients with nfvPPA, PSP and CBS relative to those with svPPA and controls. The speech of patients with lvPPA and svPPA contained a disproportionately greater number of words of both high frequency and high semantic diversity. Results from our exploratory voxel-based morphometry analyses across the whole group revealed correlations between grey matter volume in (i) bilateral frontal lobes with overall language output, (ii) the left frontal and superior temporal regions with speech complexity, (iii) bilateral frontotemporal regions with phonology, and (iv) bilateral cingulate and subcortical regions with age of acquisition. With the 15-word checklists, the LASSO models showed excellent accuracy for within-sample *k*-fold classification (over 93%) and out-of-sample validation between patients and controls (over 90%), and moderately good (59% - 74%) differentiation between the motor disorders (nfvPPA, PSP, CBS) and lexico-semantic groups (svPPA, lvPPA). In conclusion, we propose that a simple 15-word checklist provides a suitable screening test to identify people with progressive aphasia, while further specialist assessment is needed to differentiate accurately some groups (e.g., svPPA versus lvPPA and PSP versus nfvPPA).

## 1. Introduction

Speech is an integral part of effective communication and is often disturbed by brain damage such as stroke or neurodegeneration. Breakdown in speech production is important clinically as it can be diagnostic for different types of aphasia. Clinicians use conversations and narratives to detect communication difficulties in people with a speech and/or language impairment. Connected speech elicited by a picture description task, in particular, has been used to distinguish healthy controls from patients with diverse neurodegenerative diseases, as well as between specific subtypes of stroke aphasia and Primary Progressive Aphasia (PPA).^1–3^ To aid differential diagnosis and improve understanding about the nature of speech and language changes in PPA, many speech and linguistic measures have been previously investigated (e.g., acoustic/prosodic, lexico-semantic, morpho-syntactic, pragmatic/discourse) and subsequently quantified (e.g., speech rate, syllable duration, words per minute, psycholinguistic word properties) in connected speech analyses. However, transcription and quantification of speech properties require advanced linguistic expertise and are time-consuming. A simple analytical tool for analysing connected speech would be of great benefit. For example, if a simple target word list can be used (validated by in-depth, systematic analysis of connected speech with high diagnostic differentiation between progressive aphasias), this could be a practical and efficient clinical tool for assessing and diagnosing people with a neurodegenerative language impairment.

An important first step to this objective is to determine the distribution of words produced by each patient group and consider the variety of speech features and psycholinguistic properties. Both qualitative and quantitative differences in connected speech have been reported in PPA. For example, the number of content words is reduced in patients with the semantic variant (svPPA), with over-reliance on highly frequent words; in other words, the content of their speech becomes “lighter” with overuse of words that are more frequent, less concrete, less imageable, and more semantically diverse.^4,5^ Even though relatively less is known about the psycholinguistic properties of words produced by the non-fluent (nfvPPA) and logopenic (lvPPA) variants, articulatory and prosodic features, such as syllable duration, speech rate and word length, and grammatical complexity have been reported to differentiate between these two variants.^6–8^

Language impairments are also common in progressive supranuclear palsy (PSP) and corticobasal syndrome (CBS). Both conditions have features that overlap with nfvPPA^9,10^ such as dysfluency and syntactic impairments in production and comprehension.^11^ Similarities across these three groups have been reported in acoustic and lexical measures of connected speech during a picture description task.^12^ Connected speech alterations have been found in PSP patients^13–15^ including reduced speech rate, reduced total number of words and sentences, higher number of pronouns, and impaired grammatical complexity.^16,17^ Only a few studies have investigated connected speech in CBS, with one describing an overall reduction in connectedness (i.e., the number of connected events as a proportion of mentioned events) during a narrative discourse^18^ and another reporting reduced speech production rate and lexical-semantic errors during a picture description task.^19^

The differing methods of connected speech analysis in previous investigations pose a challenge in determining which measures, amongst an exhaustive list of word properties and features related to speech/language quantification, are useful for distinguishing between neurodegenerative diseases with a primary or associated language impairment. Here, we sought to address this knowledge gap with the following aims: 1) to determine which speech-related properties differentiate between svPPA, lvPPA, nfvPPA, PSP, CBS, and healthy controls during picture description using a principal component analysis to understand and simplify the patterns of change in quantifiable speech and psycholinguistic properties of connected speech; 2) to examine the neural correlates of connected speech in these conditions; and 3) to use a data-driven approach to develop an easy-to-use and practical word checklist.

## 2. Materials and methods

### 2.1 Participants

Seventy-four people (24 healthy controls, nine svPPA, nine lvPPA, nine nfvPPA, 10 PSP, 13 CBS) from the Mini Linguistic State Examination (MLSE)^20^ study were included in the development dataset. Controls were recruited through the National Institute for Health Research “Join Dementia Research” register and via local advertisement; other participants were recruited from tertiary referral services at Addenbrooke’s Hospital, Cambridge (N = 46), and Salford Royal Foundation Trust and its associated clinical providers (N = 4). Patients from a second site in the MLSE Study^20^ at St. George’s Hospital, London made an out-of-sample test set with svPPA (N = 7), lvPPA (N = 13), nfvPPA (N = 5), PSP (N = 2), and CBS (N = 6). Clinical diagnoses of PPA, PSP, and CBS were based on current consensus criteria.^21–23^ In our development sample, one nfvPPA patient declared a native language of Italian. Two svPPA patients from our out-of-sample site declared a native language of Gujarati and Indian Patois. All three patients who declared a non-English native language were pre-morbidly highly fluent in English.

### 2.2 Connected speech acquisition, transcription, reliability, and analysis

Participants completed the MLSE and the Boston Diagnostic Aphasia Examination (BDAE)^24^ and were asked to describe both the BDAE ‘cookie theft’ and MLSE ‘beach scene’ pictures. The instruction for both pictures were as follows: “Look carefully at this picture and describe aloud what is happening. Try to use sentences. I will stop you after one minute. Ready?” The examiners politely allowed time for the participants to finish their description after the one-minute mark. Connected speech samples were video recorded and transcribed by a speech-language pathologist (SKH), blinded to the clinical diagnoses, using the f4transcript notation software version 7.0, which has been previously reported to make the manual writing of speech samples from audio or video recordings more efficient. Speech samples were formatted for analysis with the Frequency in Language Analysis Tool^25^ which has specific codes for false speech. For example, false starts, grammatical errors, grammatical clause boundaries, prosodic indicators, non-lexical interjections such as filler words and pauses, repetitions, unintelligible segments, and neologisms were coded and excluded from the analyses.

To assess transcription reliability, we randomly selected 2 transcripts from each diagnostic group. We assessed the reliability of two different transcribers (SKH and KAP) by dividing the number of matching words between the two transcripts by the total words in the transcript used in our analyses (transcribed by the first author, SKH). This method is consistent with previously reported reliability analyses of transcriptions.^26^ We found a high percent agreement between the two transcripts with an average of 92% (range 81% to 98%) and 100% for the words in the 15-item checklist (below).

Using the transcribed speech samples free of false speech, we calculated the simplest measurements of connected speech (i.e., word counts, ratios, timing) to test whether these can differentiate groups as well as other measures of connected speech that tend to be more time-consuming to score and analyse (e.g., acoustic features). The total number and type counts for words, total time and words per minute were calculated for each participant. Additionally, the number and type counts for word bigrams (i.e., two word combinations such as “the mother”) and word trigrams (i.e., three word combinations such as “sink is overflowing”), type-to-token ratios for words, word bigrams, and word trigrams, proportion of function relative to content words, and combination ratio (i.e., a measure of connected language calculated as word trigram count divided by word count)^27^ were extracted using an automated script for language quantification with the use of FLAT.^25^ These measures of speech fluency for the two picture description tasks were included in our first principal component analysis (PCA).

Next, for the psycholinguistic word properties, each distinct word produced across all participants was extracted for analysis. We then excluded function words (e.g., articles, demonstratives, prepositions) and, for each content word, we looked up the ratings from various databases for length, log frequency,^28^ semantic diversity,^29^ semantic neighborhood density,^30^ concreteness,^31^ age of acquisition,^32^ orthographic and phonological Levenshtein distance.^33,34^ Where ratings for pluralised words were unavailable, word properties for the singular version were extracted. Although ratings for familiarity and imageability were initially obtained, these measures were excluded in the main analysis due to the unavailability of ratings for a high proportion of words. Of the available data, imageability ratings were strongly correlated with concreteness ratings (*R* = 0.94, *p* < 0.001) and familiarity ratings were moderately correlated with log frequency ratings (*R* = 0.45, *p* < 0.001). These word properties were included in our second PCA.

### 2.3 Statistical analysis

We used PCA as a dimensionality reduction method to investigate the distinct speech characteristics underlying connected speech performance. First, we calculated the average counts per participant for the quantifiable measures of speech fluency (e.g., number and type of words, type to token ratio, word per minute) using the transcribed speech samples which were then entered into a varimax-rotated PCA. A Kaiser-Meyer-Olkin test determined the suitability of our dataset. We selected three components based on Cattell’s criterion. Using principal component scores per participant, we conducted a one-way analysis of variance (ANOVA) to test for group differences.

Next, to understand the underlying pattern of variations in the lexico-semantic word properties produced by all patients and controls, all unique content words produced by patients and controls in both picture descriptions were compiled into a single ‘speech corpus’ and the psycholinguistic properties of each word were entered into a varimax-rotated PCA. After selecting three components using Cattell’s criteria, principal component scores for the words produced by each participant were extracted and then averaged across individual participants. Using these averaged principal component scores per participant, we tested the differences between group and task (i.e., ‘cookie theft’ versus ‘beach scene’) using a two-way ANOVA. Past studies have found that comparison of mean values can be relatively insensitive for detecting patients’ altered word usage, whereas distribution analyses can be more sensitive (e.g., where there are more pronounced changes in one part of the distribution).^4,5^ Thus, using data from the word properties PCA, principal component scores were split into quartiles (ranging from −4 to 2-, greater than −2 to 0, greater than 0 to 2, and greater than 2 to 4). For each participant, we counted the number of times each participant produced words in each range of a principal component (e.g., −4 to −2 in PC 1) and each point in the psycholinguistic dimensional space (e.g., −4 to −2 in PC 1 and 2 to 4 in PC 2). We then generated contour plots that mapped the proportion of words produced by each participant which were then averaged across groups. Using a method previously applied by Hoffman *et al*.^5^ we generated difference plots by subtracting the mean of control data from that of each patient group’s data to visualise the differences between control versus patient maps. We explored differences between groups across the variation in word properties in two ways. First, we took the mean value of the proportion of words produced by each patient group and compared them to the control data in each of the dimensional spaces using two-tailed *t*-tests. Secondly, for a more sensitive method, we conducted a distribution analysis by quantifying the number of words produced by controls and patients in each of the principal components’ quartiles. A repeated measures ANOVA was performed with quartiles as within-subject and group as between-subject factors.

*Post hoc* analyses were conducted using Tukey’s honestly significant difference (HSD) test for multiple comparison. All statistical analyses were performed in R statistical software (version 2023.03.0).

### 2.4 Neuroimaging acquisition and voxel-based morphometry analysis

All participants underwent T_1_-weighted structural MRI of the brain. Participants from Cambridge were scanned using a 3T Siemens Skyra MRI scanner. Whole-brain T_1_-weighted structural images were acquired using the following parameters: iPAT2; 208 contiguous sagittal slices; field of view (FOV) = 282 x 282 mm^2^; matrix size 256 x 256; voxel resolution = 1.1 mm^3^; TR/TE/ TI = 2000 ms/2.93 ms/850 ms, respectively; and flip angle 8°. Participants from Manchester were scanned using a 3T Philips Achieva MRI scanner. Whole-brain T_1_-weighted images were acquired using the following parameters: SENSE = 208 contiguous sagittal slices; FOV = 282 x 282 mm^2^; matrix size 256 x 256; voxel resolution = 1.1mm^3^; TR/TE/TI = 6600 ms/2.99 ms/850 ms, and flip angle 8°.

Whole-brain grey matter changes were indexed using voxel-based morphometry (VBM) analyses of structural T_1_-weighted MRI, integrated into Statistical Parametric Mapping software (SPM12: Wellcome Trust Centre for Neuroimaging, https://www.fil.ion.ucl.ac.uk/spm/software/spm12/). A standard pre-processing pipeline was implemented involving: (i) brain segmentation into three tissue probability maps (grey matter, white matter, cerebrospinal fluid); (ii) normalisation (using Diffeomorphic Anatomical Registration Through Exponentiated Lie Algebra, DARTEL);^35^ (iii) study-specific template creation using grey matter tissue probability maps; (iv) spatial transformation to Montreal Neurological Institute (MNI) space using transformation parameters from the corresponding DARTEL template; and (v) image modulation and smoothing using 8mm full-width-half-maximum Gaussian kernel to increase signal-to-noise ratio. Segmented, normalised, modulated and smoothed grey matter images were used for VBM analyses.

We examined the associations between whole-brain grey matter intensity and PCA-generated principal component scores, which were averaged across two picture description tasks, using *t*-contrasts. Age and total intracranial volume were included as nuisance covariates. Clusters were extracted using a threshold of *p* < 0.001 uncorrected for multiple comparisons with a cluster threshold of 100 voxels. We chose 100 voxels as our cluster threshold as we were interested in smaller subcortical regions that have been reported to be associated with speech production and are often atrophic in the disease groups.

### 2.5 Word checklist analysis

To determine target words that could best differentiate between groups, we used Least Absolute Shrinkage and Selection Operation (LASSO) logistic regression.^36^ Given the large number of predictors (i.e., 500+ unique words used by the whole group), relatively small sample size per group, and multicollinearity of the words (e.g., the likelihood that a participant would say “overflowing” and “sink”), the LASSO method is highly appropriate for automated feature selection and shrinkage. While multiple correlated words are entered into the model, only the most important predictor variables (i.e., the least number of words that best differentiate between groups) will be selected. As a first step, we pooled together all of the different words that patients and controls produced which resulted in over 500+ tokens per picture. We then streamlined this collection by carrying out LASSO regressions for each picture including all unique words produced per picture as predictors for the following comparisons: (i) controls versus each patient group, and (ii) each patient group against one another. Whether or not a participant produced a word such as “overflowing” was coded as 1 for produced and 0 for not produced. We accounted for differences in dialect (e.g., score of 1 if the participant said boy, chap, lad, or bloke) and morpho-syntax such as verb tense (e.g., stealing/stolen) and singular/plural forms (e.g., plate/plates).

Next, the words that had been selected in each pairwise comparison (by logistic LASSO regression) were compiled, resulting in a pool of 33 words for the BDAE ‘cookie theft’ and 46 words for the MLSE ‘beach scene’ pictures. We re-ran the LASSO regressions for each pairwise comparison using these truncated lists and the resulting words were further rank ordered by (i) the number of times they appear in the pairwise comparisons, (ii) their beta coefficients, and (iii) the magnitude of difference in the overall proportion by group (e.g., magnitude would be 1 if all of the controls produced the word ‘overflow’ but none of the svPPAs did). The top 15 words resulting from this rank ordering were entered into a series of four-fold cross-validated LASSO logistic regressions with each predicting the diagnostic distinction of interest (e.g., controls versus patients). The scoresheets using the 15 words are shown in Appendix 1.

To evaluate the robustness of the model in predicting group classification with the word checklists, we conducted out-of-sample predictive validity testing with connected speech data from St. George’s Hospital. There were no differences in demographics between patients from the two test sites except for PSP patients from St. George’s having lower scores on the revised Addenbrooke’s Cognitive Examination (ACE-R) compared to those from Cambridge (*p* = 0.02). We tested the 15-word checklist with the St. George’s data assigning a score of 1 if the participant produced the target word and a 0 if the word was omitted. Morpho-syntactic variations were scored as correct if the root matched the target word (e.g., overflowing for overflow, digging for dig). As an index of accuracy for our binomial models (i.e., pairwise comparisons), we report classification performance on the test data using function confusion.glmnet from the glmnet package in R for the following comparisons: controls versus all patients, patients belonging to the “motor” group (i.e., nfvPPA, PSP, CBS) versus “lexico-semantic” group (i.e., svPPA, lvPPA), and each patient group against one another. Of note, PSP and CBS patients were grouped into one due to small sample size (i.e., 2 PSP) in our out-of-sample test set.

To test the hypothesis that supplementing the checklist with cognitive scores might improve the differentiation between groups, we ran another LASSO logistic regression with the 15 words (coded the same way as noted above), as well as subtest scores from the ACE-R and MLSE. We estimated the LASSO model using a within-sample four-fold cross-validation with the Cambridge training set and tested the generalisability of our model with the St. George’s data as out-of-sample test.

## 3. Results

### 3.1 Demographics

Demographic and clinical features are shown in Table 1. There were no significant differences in all groups for age, gender, and handedness, as well as symptom duration for patients. There were significant differences between groups in education; *post hoc* tests confirmed that controls left education later than patients with nfvPPA, CBS, and PSP (*p* < 0.05). Significant group differences emerged on total MLSE and ACE-R scores. Controls performed better on the MLSE when compared with patients with svPPA, lvPPA, nfvPPA, and CBS (*p* < 0.001), PSP performed better than lvPPA (*p* = 0.001) and nfvPPA (*p* = 0.007), and CBS performed better than lvPPA (*p* = 0.03). On the ACE-R, controls performed better than all patient groups (*p* < 0.05), and nfvPPA, PSP, and CBS performed better than lvPPA (*p* < 0.05), and PSP performed better than svPPA (*p* = 0.001). As shown in Table 1, in our development sample, all participants were white. Two svPPA patients from our out-of-sample site were non-white.

**Table 1.** Demographics and clinical features of the study cohort.

| | Control | svPPA | lvPPA | nvPPA | PSP | CBS | $p^*$ |
| --- | --- | --- | --- | --- | --- | --- | --- |
| N | 24 | 9 | 9 | 9 | 10 | 13 | - |
| Age (SD) | 65.8 (5.2) | 67.2 (4.3) | 68.9 (8.1) | 70.1 (6.4) | 68.4 (5.9) | 70.2 (4.4) | ns |
| Gender M:F | 11:13 | 5:4 | 6:3 | 4:5 | 5:5 | 7:6 | ns |
| Handed-ness R:L | 21:3 | 9:0 | 9:0 | 8:1 | 9:1 | 12:1 | ns |
| Ethnicity<br>White:Other | 24:0 | 9:0 | 9:0 | 9:0 | 9:0 | 9:0 | - |
| First language<br>English:Other | 24:0 | 9:0 | 9:0 | 8:1 | 9:0 | 9:0 | - |
| Age left education<br>(SD) | 20.6 (3.3) | 19.3 (2.6) | 20.6 (4.1) | 16.6 (1.7) | 16.7 (1.7) | 17.4 (3.0) | < 0.001 |
| Symptom duration<br>in years (SD) | NA | 6.5 (2.5) | 3.0 (2.7) | 3.2 (2.9) | 4.1 (2.5) | 5.2 (4.0) | ns |
| Total MLSE (SD) | 98.3 (2.2) | 78.1 (4.7) | 68.1 (15.3) | 70.9 (15.5) | 87.9 (8.0) | 81.8 (14.3) | < 0.001 |
| ACE-R (SD) | 96.0 (3.4) | 53.9 (8.2) | 46.7 (25.1) | 69.7 (15.1) | 80.5 (13.4) | 74.0 (17.6) | < 0.001 |

Note: Mean and standard deviations are displayed. For MLSE and ACE-R, values indicate scores out of 100. \**p*-value for F-test of group-difference by ANOVA. ACE-R, Addenbrooke's Cognitive Examination Revised; CBS, corticobasal syndrome; lvPPA, logopenic variant primary progressive aphasia; MLSE, Mini Linguistic State Examination; nfvPPA, non-fluent variant primary progressive aphasia; ns, not significant, $p > 0.1$ ; PSP, progressive supranuclear palsy; SD, standard deviation; svPPA, semantic variant primary progressive aphasia.

### 3.2 Quantification of speech fluency

Average counts per participant for the quantifiable properties of words and word combinations per picture were entered into a PCA with varimax rotation. Three principal components were identified using Cattell’s criteria which explained 86.5% of the variance (Kaiser-Meyer-Olkin = 0.70). The loadings of each measure are shown in Supplementary Table 1.

Type and token counts for words, word bigrams, and word trigrams, word per minute, type-to-token ratio of words, and combination ratio loaded most heavily on principal component (PC) 1 and thus we labelled this PC as ‘speech quanta’. Type-to-token ratio of words, word bigrams, and word trigrams loaded most heavily on PC 2 which we labelled as ‘lexical richness’. Word per minute, an index of speech fluency, and combination ratio, the degree to which an individual produced longer, more-complex combinations as opposed to single word fragments, loaded heavily on PC 3 and we adopted the working label of ‘speech complexity’.

Group performance patterns on all three PCs are visually summarised in Figure 1A. For PC 1, the results from a one-way ANOVA revealed group differences (F(1,142) = 71.19, *p* < 0.001), driven by controls and svPPA patients having higher scores than those with nfvPPA (*p* < 0.001), PSP (*p* < 0.01), and CBS (*p* < 0.05). Additionally, controls had higher scores than patients with lvPPA (*p* = 0.01), who in turn had higher scores than those with nfvPPA (*p* < 0.001). A one-way ANOVA did not reveal group differences for PC 2 (F(1,142) = 1.26, *p* = 0.26). For PC 3, the results from a one-way ANOVA revealed group differences (F(1,142) = 12.77, *p* < 0.001), driven by controls having higher scores than those with nfvPPA (*p* < 0.001), PSP (*p* < 0.001), and CBS (*p* = 0.002).

**Figure 1.**
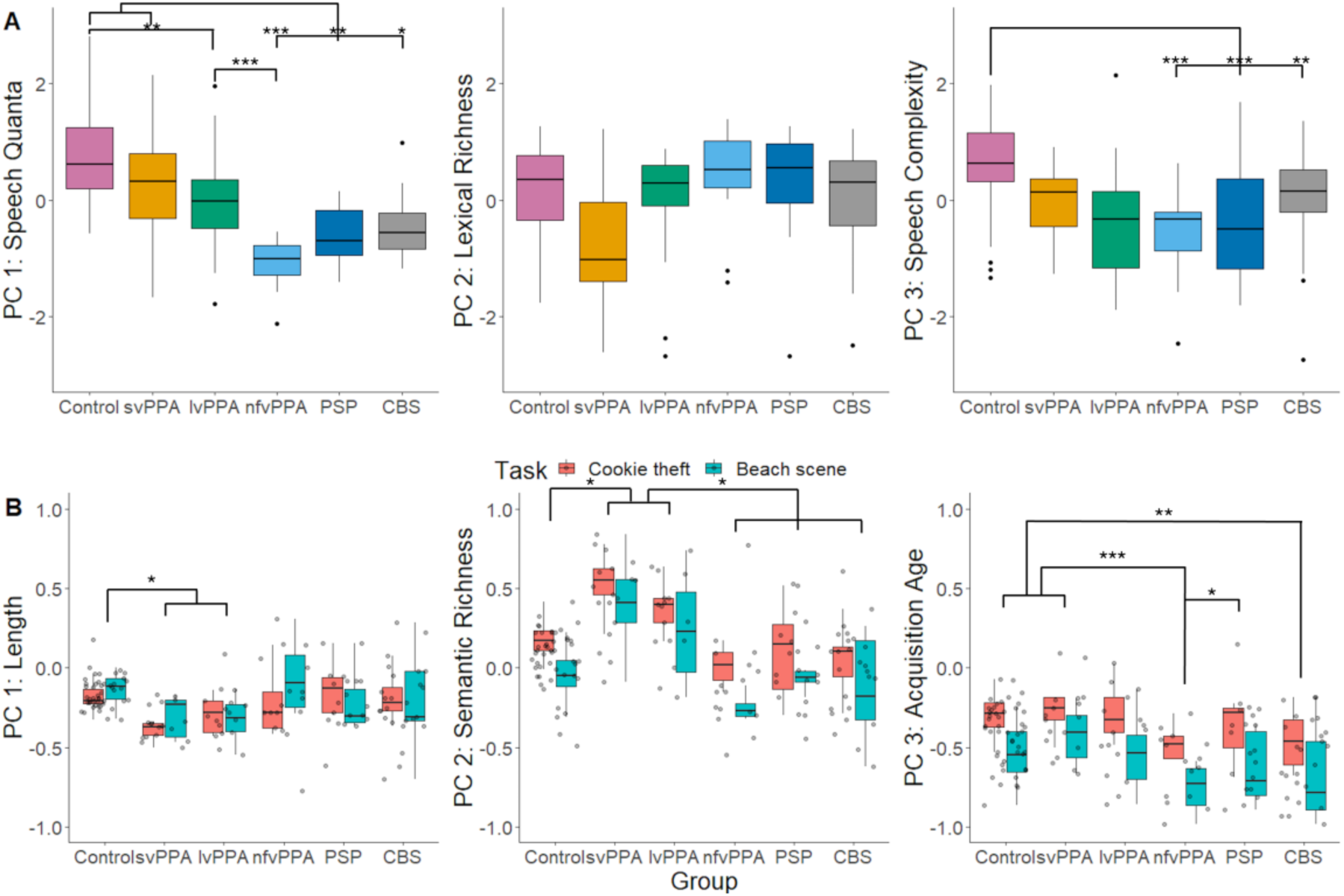
Principal component analysis scores across diagnostic groups. (**A**) Scores of quantitative measures of speech fluency. For PC 1 (‘speech quanta’), the results from a one-way ANOVA revealed significant group differences (F(1,142) = 71.19, *p* < 0.001), driven by controls (N = 24) and patients with svPPA (N = 9) having higher scores than those with nfvPPA (N = 9), PSP (N = 10), and CBS (N = 13), and controls having higher scores than those with lvPPA (N = 9), and patients with lvPPA having higher scores than those with nfvPPA. PC 2 (‘lexical richness’) resulted in no group differences (F(1,142) = 1.26, *p* = 0.26), and for PC 3 (‘speech complexity’) significant group differences were found (F(1,142) = 12.77, *p* < 0.001), driven by controls having higher scores than patients with nfvPPA (*p* < 0.001), PSP (*p* < 0.001), and CBS (*p* = 0.002). (**B**) Scores of quantitative measures of word properties across groups. For PC 1 (‘length’), the results from a two way ANOVA revealed significant group differences (F(5,134) = 4.29, *p* < 0.001), driven by svPPA and lvPPA patients producing words that were shorter, phonologically and orthographically less complex than controls (*p* < 0.05). For PC 2 (‘semantic richness’), significant differences were found for group (F(5,134) = 16.62, *p* < 0.001) and task (F(1,134) = 22.05, *p* < 0.001). Patients with svPPA and lvPPA produced more words that were characterised as more frequent and semantically diverse than those with nfvPPA, PSP, CBS, and controls (*p* < 0.01). For PC 3 (‘acquisition age’), significant differences were found for group (F(5,134) = 7.09, *p* < 0.001) and task (F(1,134) = 50.24, *p* < 0.001) for PC 3. *Post hoc* analyses revealed that (i) nfvPPA patients produced words that were characterised as significantly earlier acquired than those with svPPA (*p* < 0.001), PSP (*p* = 0.05), and controls (*p* < 0.001), and (ii) CBS patients used words that were significantly earlier acquired than those with svPPA (*p* = 0.002) and controls (*p* = 0.01). Results from *post hoc* analyses using Tukey’s honestly significant difference (HSD) test for multiple comparisons are shown as asterisks indicating level of significance: * *p* ≤ 0.05; ** *p* ≤ 0.01; *** *p* ≤ 0. 001. CBS, corticobasal syndrome; lvPPA, logopenic variant of primary progressive aphasia; nfvPPA, non-fluent variant of primary progressive aphasia; PC, principal component; PSP, progressive supranuclear palsy; svPPA, semantic variant of primary progressive aphasia.

Correlations between the speech fluency PC scores and total and subdomain scores of the MLSE can be found in Supplementary Table 2.

### 3.3 Quantification of word properties

Ratings of psycholinguistic features for all words produced by controls and patients were entered into a PCA with varimax rotation. Three principal components were identified using Cattell’s criteria, each representing a group of covarying psycholinguistic features. These three components explained 85.5% of the variance (Kaiser-Meyer-Olkin = 0.75). The loadings of each measure are shown in Supplementary Table 3. Length, phonological and orthographic Levenshtein distance loaded most heavily on PC 1 and we adopted the working label of ‘length’. Concreteness, log frequency, semantic neighbourhood density, and semantic diversity loaded heavily on PC 2 which we labelled as ‘semantic richness’. Age of acquisition loaded most heavily on PC 3 which we labelled as ‘acquisition age’.

The three scores, obtained from the psycholinguistic PCA results, per participant along with the elicitation task were into a two-way ANOVA which revealed significant group differences in PC 1 (F(5,134) = 4.29, *p* < 0.001), driven by svPPA and lvPPA patients producing words that were shorter, phonologically and orthographically less complex than controls (*p* < 0.05) (see Figure 1B).

For PC 2, significant differences were found for group (F(5,134) = 16.62, *p* < 0.001) and task (F(1,134) = 22.05, *p* < 0.001). The task effect was driven by more frequent and semantically diverse words produced for the ‘cookie theft’ than the ‘beach scene’ picture. *Post hoc* analyses revealed that svPPA and lvPPA patients produced more words that were characterised as more frequent and semantically diverse than those with nfvPPA, PSP, CBS, and controls (*p* < 0.01).

Significant differences were found for group (F(5,134) = 7.09, *p* < 0.001) and task (F(1,134) = 50.24, *p* < 0.001) for PC 3. The words used to describe the ‘cookie theft’ were found to be later acquired. *Post hoc* analyses revealed that nfvPPA patients produced words that were characterised as significantly earlier acquired than those with svPPA (*p* < 0.001), PSP (*p* = 0.05), and controls (*p* < 0.001). Similarly, CBS patients used words that were significantly earlier acquired than those with svPPA (*p* = 0.002) and controls (*p* = 0.01).

Correlations between the word properties PC scores and total and subdomain scores of the MLSE can be found in Supplementary Table 2.

#### 3.3.1 Differences in multivariate word properties

Moving beyond the simplistic mean statistic, we looked at the bivariate distributions of words across the psycholinguistic space and how these might shift in each patient group (e.g., patients produce fewer words in one part of the space and might substitute more words in another part of the space). Figure 2 shows the contour plot for controls (left), depicting the averaged proportion of words produced within the principal component space, and the difference plots where the mean of the control data for the three principal components from the ‘word properties’ PCA (Section 3.3) were subtracted from that of the patient data.

**Figure 2.**
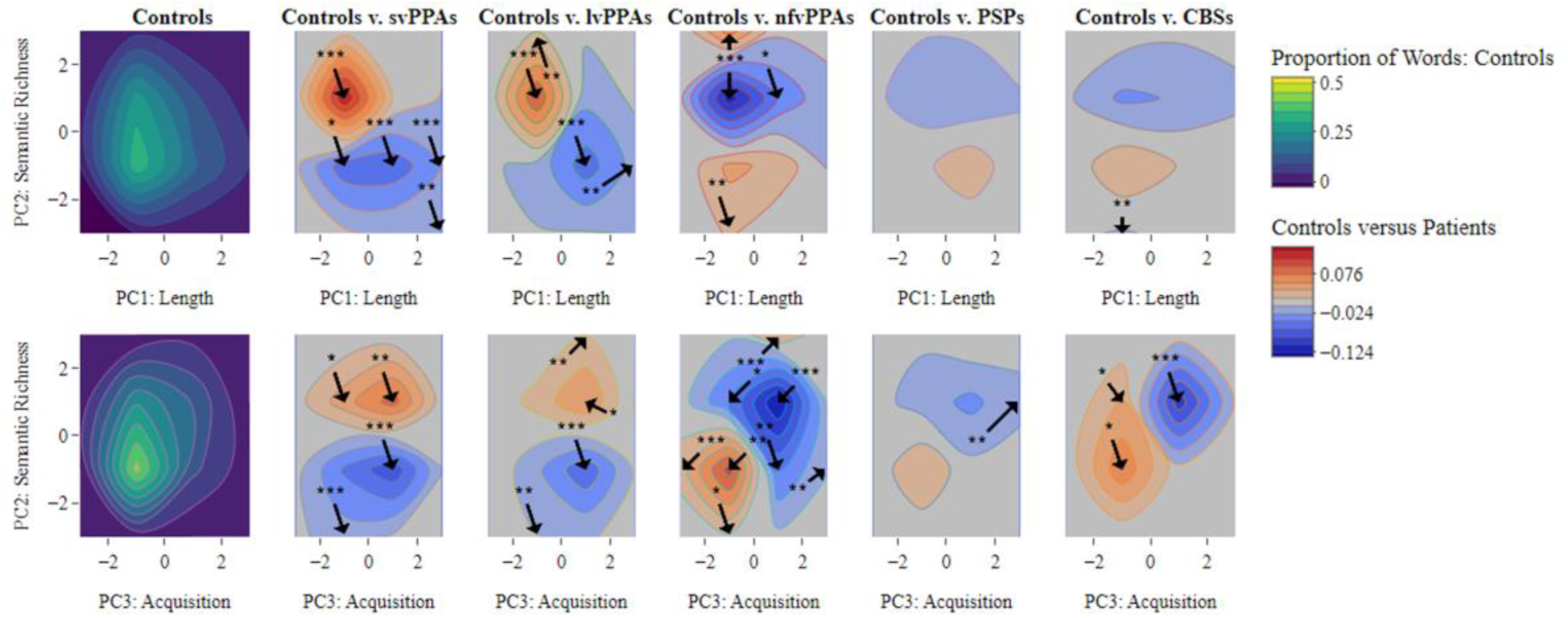
Contour distributions and difference plots. The top and bottom left plots show the contour distributions across principal component (PC) 1: Length, PC 2: Semantic richness, and PC 3: Acquisition age produced by healthy controls. Difference plots comparing patients with healthy controls are shown to the right of the contour plots of healthy controls. In the control plots, yellow tones show where the greatest proportions of words were found within the principal component space. For controls versus patients, the red and blue tones represent principal component spaces where patients produced more words than controls and where controls produced more than patients, respectively. Taking the mean value of the proportion of words produced by each patient group (svPPA N = 9, lvPPA N = 9, nfvPPA = 9, PSP N = 10, CBS N = 13), we compared them to the control data (N = 24) in each of the dimensional spaces using two-tailed *t*-tests. The arrows indicate where in the maps there were significant differences between controls and patients (*p*-values are shown as asterisks indicating level of significance: * *p* < 0.05; ** *p* < 0.01; *** *p* < 0. 001).

Relative to controls, svPPA and lvPPA patients produced a greater proportion of words in the higher semantic richness (i.e., more semantically diverse and frequent) and lower length (i.e., shorter, less phonologically and orthographically complex) space. In contrast, nfvPPA, PSP, and CBS patients produced a greater proportion of words with lower semantic richness and acquisition age (i.e., earlier acquired) space.

#### 3.3.2 Distribution analysis of word properties PCA

Another way to go beyond the simplistic mean statistic is to undertake a formal distribution analysis for each principal component. This has been shown in previous work to be much more sensitive to changes in the content words produced by patients.^37,38^ As shown in Figure 3, principal component scores for PC 1 to PC 3 from the word properties PCA were divided into quartiles and the number of words produced in each quartile was computed for each participant followed by a group mean.

**Figure 3.**
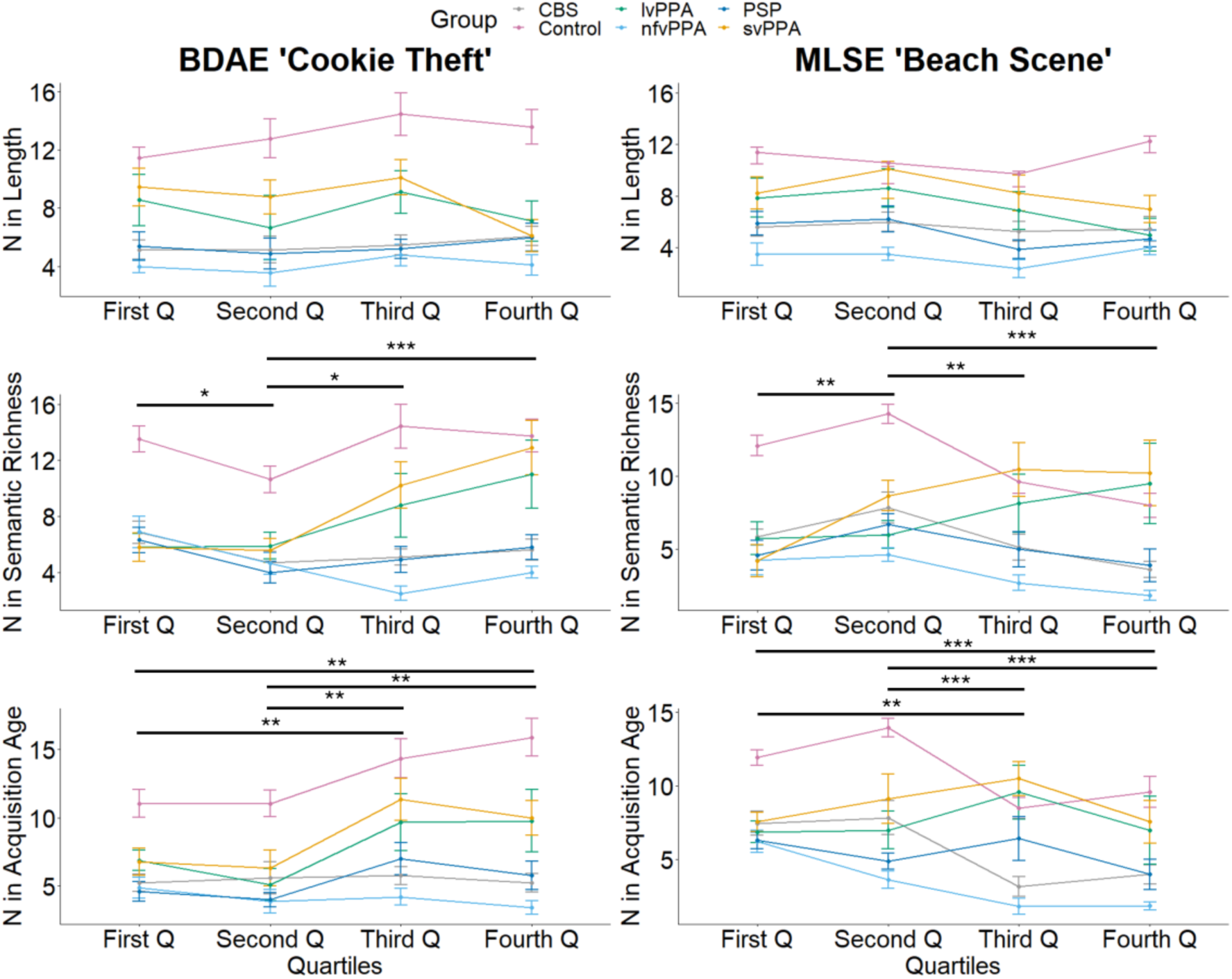
Distribution plots. Each plot shows the number (N) of words produced in each quartile (Q) by patient groups (svPPA N = 9, lvPPA N = 9, nfvPPA = 9, PSP N = 10, CBS N = 13) for principal component (PC) 1 ‘length’, PC 2 ‘semantic richness’, and PC 3 ‘acquisition age’. For PC 1, a six groups x four quartiles repeated measures ANOVA showed a significant effect of group only for both ‘cookie theft’ (F(5,283) = 37.16, *p* < 0.001) and ‘beach scene’ (F(5,272) = 39.18, *p* < 0.001). For PC 2, significant effects were found for group (F(5,280) = 33.68, *p* < 0.001), quartile (F(1,280) = 4.67, *p* = 0.03), and group-by-quartile interaction (F(5,280) = 4.36, *p* < 0.001) for ‘cookie theft’. For ‘beach scene’, significant effects were found for group (F(5,270) = 28.94, *p* < 0.001), quartile (F(1,270) = 5.53, *p* = 0.02), and group-by-quartile interaction (F(5,270) = 8.29, *p* < 0.001). For PC 3, significant effects were found for group (F(5,283) = 36.15, *p* < 0.001), quartile (F(1,283) = 17.17, *p* < 0.001), and group-by-quartile interaction (F(5,283) = 2.47, *p* = 0.03) for ‘cookie theft’. For ‘beach scene’, significant effects were found for group (F(5,265) = 31.04, *p* < 0.001), quartile (F(1,265) = 21.67, *p* < 0.001), and group-by-quartile interaction (F(5,265) = 2.47, *p* = 0.03). The effect of quartile from *post hoc* analyses using Tukey’s honestly significant difference (HSD) test for multiple comparisons is shown as asterisks indicating level of significance: * *p* ≤ 0.05; ** *p* ≤ 0.01; *** *p* ≤ 0. 001. BDAE, Boston Diagnostic Aphasia Examination; CBS, corticobasal syndrome; lvPPA, logopenic variant of primary progressive aphasia; MLSE, Mini Linguistic State Examination; nfvPPA, non-fluent variant of primary progressive aphasia; PSP, progressive supranuclear palsy; svPPA, semantic variant of primary progressive aphasia.

For PC 1, a six groups x four quartiles repeated measures ANOVA showed a significant effect of group only for both ‘cookie theft’ (F(5,283) = 37.16, *p* < 0.001) and ‘beach scene’ (F(5,272) = 39.18, *p* < 0.001). For PC 2, a six groups x four quartiles repeated measures ANOVA showed significant effects of group (F(5,280) = 33.68, *p* < 0.001), quartile (F(1,280) = 4.67, *p* = 0.03), and group-by-quartile interaction (F(5,280) = 4.36, *p* < 0.001) for ‘cookie theft’. For ‘beach scene’, a six groups x four quartiles repeated measures ANOVA showed significant effects of group (F(5,270) = 28.94, *p* < 0.001), quartile (F(1,270) = 5.53, *p* = 0.02), and group-by-quartile interaction (F(5,270) = 8.29, *p* < 0.001). For PC 3, a six groups x four quartiles repeated measures ANOVA showed significant effects of group (F(5,283) = 36.15, *p* < 0.001), quartile (F(1,283) = 17.17, *p* < 0.001), and group-by-quartile interaction (F(5,283) = 2.47, *p* = 0.03) for ‘cookie theft’. For ‘beach scene’, a six groups x four quartiles repeated measures ANOVA showed significant effects of group (F(5,265) = 31.04, *p* < 0.001), quartile (F(1,265) = 21.67, *p* < 0.001), and group-by-quartile interaction (F(5,265) = 2.47, *p* = 0.03). Our results are summarised in Supplementary Table 4.

### 3.4 Neural correlates of connected speech properties

Associations between grey matter intensity and principal component scores from both quantitative measures of speech fluency and word properties are shown in Figure 4 and Supplementary Table 5. In the entire group (i.e., patients and controls), PC 1 (‘speech quanta’) scores correlated with grey matter intensities of the bilateral middle and superior frontal gyri, right inferior frontal gyrus, insula, putamen, and caudate. PC 3 (‘speech complexity’) scores correlated with grey matter intensities of the left insula, inferior, middle, and superior frontal gyri, extending medially, superior temporal gyrus, and parts of the limbic system. No significant correlations were found for PC2 (‘lexical richness’) scores.

**Figure 4.**
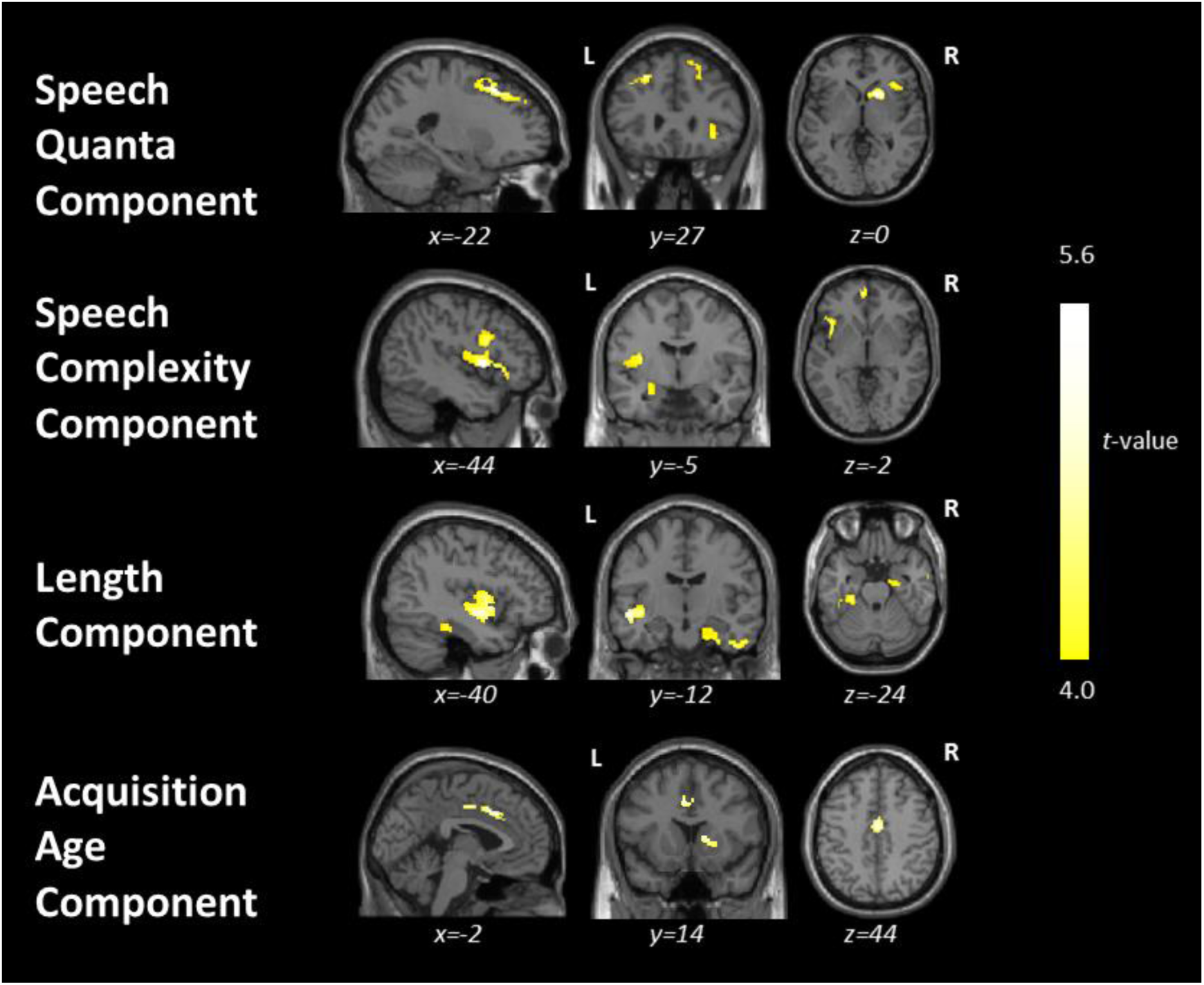
Results from the whole-brain voxel-based morphometry correlation analyses. This figure shows regions of grey matter intensity that uniquely correlate with principal component scores in the whole group including controls (N = 24) and patients (svPPA N = 9, lvPPA N = 9, nfvPPA = 9, PSP N = 10, CBS N = 13) using *t*-contrasts. Clusters were extracted using a threshold of *p* < 0.001 uncorrected for multiple comparisons with a cluster threshold of 100 voxels with age and total intracranial volume included as nuisance covariates. CBS, corticobasal syndrome; lvPPA, logopenic variant of primary progressive aphasia; nfvPPA, non-fluent variant of primary progressive aphasia; PSP, progressive supranuclear palsy; svPPA, semantic variant of primary progressive aphasia.

For the word properties PCA, PC 1 (‘length’) scores correlated with grey matter intensities of the left insula, middle and superior temporal gyri, bilateral parahippocampal and fusiform gyri, right inferior and middle temporal gyri, and limbic structures. PC 3 (‘acquisition age’) scores correlated with grey matter intensities of the bilateral cingulate gyri and right caudate and putamen. No significant correlations were found for PC 2 (‘semantic richness’) scores.

When excluding healthy controls, PC 1 (‘length’) scores correlated significantly with a single cluster including the left insula, middle and superior temporal gyri (see Supplementary Table 6). No significant correlations were found for the other PC scores. Supplementary Table 7 shows the results when using a cluster-forming height threshold of *p* < 0.005 paired with a cluster extent threshold of *p* < 0.05 FWE-corrected.

### 3.5 Word checklist

Using the word checklist for each picture (Appendix 1), the LASSO logistic regression selected a group of words that together predicted group membership (see Supplementary Table 8). Of note, the LASSO regression for svPPA versus lvPPA, and nfvPPA versus PSP resulted in zero words for both pictures; in other words, none of these words could differentiate between these groups. These results motivated our hierarchical classification as shown in Figure 5, where the “motor” group included patients with nfvPPA, PSP, and CBS, and the “lexico-semantic” group included those with svPPA and lvPPA. The within-sample *k*-fold validation accuracies for ‘cookie theft’ were as follows: 96% for patients versus controls and 92% for “motor” versus “lexico-semantic” groups. Out-of-sample test accuracy with the St. George’s data (N = 34) resulted in 91% for patients versus controls and 74% for “motor” versus “lexico-semantic” groups.

**Figure 5.**
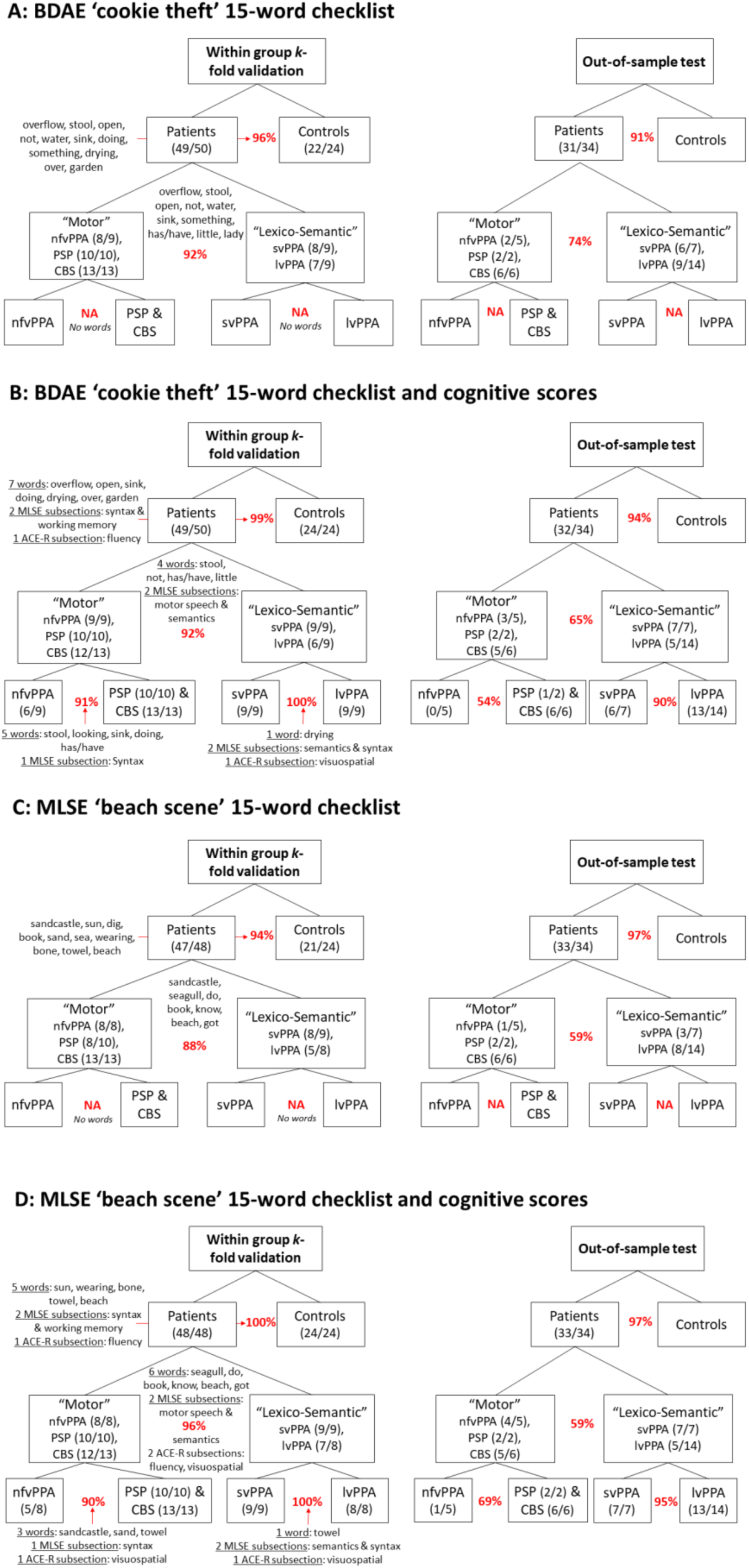
Within-sample *k*-fold and out-of-sample validations. This figure summarises the validations for (**A**) BDAE ‘cookie theft’ 15-word checklist, (**B**) BDAE ‘cookie theft’ 15-word checklist with cognitive measures of ACE-R and MLSE, (**C**) MLSE ‘beach scene’ 15-word checklist, and (**D**) MLSE ‘beach scene’ 15-word checklist with cognitive measures of ACE-R and MLSE. The curved brackets under the group names (e.g., Patients, “Motor”, “Lexico-Semantic”) illustrate the number of participants who were classified correctly out of the total N. The percentages in red indicate the within-sample four-fold and out-of-sample validation accuracies. ACE-R, Addenbrooke’s Cognitive Examination – Revised; BDAE, Boston Diagnostic Aphasia Examination; CBS, corticobasal syndrome; lvPPA, logopenic variant of primary progressive aphasia; MLSE, Mini Linguistic State Examination; nfvPPA, non-fluent variant of primary progressive aphasia; PSP, progressive supranuclear palsy; svPPA, semantic variant of primary progressive aphasia.

For ‘beach scene’, the within-sample *k*-fold validation accuracies were as follows: 94% for patients versus controls and 88% for “motor” versus “lexico-semantic” groups. Out-of-sample test accuracy resulted in 97% for patients versus controls and 59% for “motor” versus “lexico-semantic” groups. Of note, the LASSO regression for nfvPPA versus PSP and CBS combined also resulted in zero words for both pictures.

Since we were not able to differentiate individual patient groups using the checklist alone, we tested the hypothesis that supplementing with cognitive measures might improve the differentiation between these groups. To this end, we supplemented the LASSO models with ACE-R and MLSE sub-scores along with the target words and found improved differentiation for within-sample validation for both nfvPPA versus PSP and CBS (91% for ‘cookie theft’ and 90% for ‘beach scene’) and svPPA versus lvPPA groups (100% for both ‘cookie theft’ and ‘beach scene’). Moreover, results from the out-of-sample predictive validity testing showed that the checklists and LASSO models were generalisable more for svPPA versus lvPPA (90% for ‘cookie theft’ and 95% for ‘beach scene’) when compared with nfvPPA versus PSP and CBS (54% for ‘cookie theft’ and 69% for ‘beach scene’).

In Figure 5, the curved brackets under the group names (e.g., Patients, “Motor”, “Lexico-Semantic”) illustrate the number of participants who were classified correctly. Supplementary Figure 1 shows each participant’s scores on the 15-word checklists and ACE-R and highlights the participants who were misclassified. The sensitivity and specificity of the word checklists are shown in Supplementary Figure 2.

## 4. Discussion

Clinical impressions from listening to patients’ speech are often used to guide diagnosis but there are two main challenges that this study addresses. First, it is not clear what aspects of the speech should be the target of the assessment. Second, although samples of speech are easy to collect, detailed analyses of connected speech are time-consuming and require specialist expertise. In the present study, we undertook detailed transcription and analyses of connected speech elicited by two picture description tasks and established which speech features and/or psycholinguistic properties might show the greatest differentiation across groups. We then identified the atrophy correlates of speech-related features. Finally, using data-driven methods, we established a clinically efficient and effective vocabulary checklist method to aid differential diagnosis between the subtypes of primary progressive aphasia (PPA), progressive supranuclear palsy (PSP), and corticobasal syndrome (CBS).

We found significant differences in both speech features and psycholinguistic properties of words between patients and controls. These features also differentiated svPPA and lvPPA versus the remaining groups which are most typically associated with a tauopathy and/or motor disorders (nfvPPA, CBS, PSP). The total language output was significantly reduced in patients with nfvPPA, PSP, and CBS relative to those with svPPA and controls. Inspection of the proportion of words produced across the lexico-semantic space revealed that patients with svPPA and lvPPA used a greater proportion of words with high semantic richness (i.e., more frequent and semantically diverse) and lower length (i.e., shorter, less phonologically and orthographically complex) such as “do”, “out”, and “get” relative to controls. In contrast, patients with nfvPPA, PSP, and CBS showed the opposite pattern with a greater proportion of words in the lower semantic richness and acquisition age (i.e., earlier acquired) space such as “dog”, “boy”, and “cookie”.

We demonstrated that a straightforward word checklist can provide a “user-friendly” tool, quantifiable in a simple way, with high sensitivity in differentiating healthy controls from patients with a progressive aphasia. The 15-word checklist showed excellent accuracy for within-sample *k*-fold validation, for differentiating patient groups from controls. Even on an out-of-sample validation dataset, the 15-word checklist was excellent at differentiating patients from controls (out-of-sample test accuracy of 91% and 97% for ‘cookie theft’ and ‘beach scene’) and moderately good at differentiating primary “lexico-semantic” (svPPA, lvPPA) from “motor” (nfvPPA, PSP, CBS) groups (accuracy of 74% and 59% for ‘cookie theft’ and ‘beach scene’). The 15 words did not accurately differentiate patients with svPPA from lvPPA, or nfvPPA from PSP and CBS. This is perhaps unsurprising given the patients’ similar patterns of word usage, total language output, and psycholinguistic properties of the words elicited. Supplementing the 15-word checklist with cognitive measures of ACE-R and MLSE subtest scores increased diagnostic accuracy for nfvPPA versus PSP and CBS for within-sample validation (91% for ‘cookie theft’ and 90% for ‘beach scene’), as well as svPPA versus lvPPA for both within-sample (100% for both ‘cookie theft’ and ‘beach scene’) and out-of-sample validation (90% for ‘cookie theft’ and 95% for ‘beach scene’). With regard to differentiating patients from controls, the best ACE-R subtest was verbal fluency which replicates a recent study that found this simple clinical assessment is excellent at differentiating patients from controls but has limited use for differential diagnosis between patient subgroups.^39^ We propose that the quick and simple 15-word checklist is a suitable screening test to identify people with progressive aphasia, although further specialist assessment is needed for accurate diagnostic sub-typing. In the following sections, we interpret these findings, consider their clinical implications, and note directions for future research.

### Reduced language output from nfvPPA, PSP and CBS

Patients with nfvPPA, PSP and CBS were distinguishable from those with svPPA, lvPPA and controls, based on reduced language output and connected speech fluency (as measured by the ‘speech quanta’ and ‘speech complexity’ PCs). In particular, combination ratio has been previously proposed as a measure of connected language output because it represents the degree to which an individual produces longer, more-complex combinations of words over the total word count.^27^ Many studies have suggested that measures such as reduced language output, slowed articulation rate, speech-sound errors, and proportion of function to content words, can differentiate patients with nfvPPA from the other variants of PPA in connected speech and other language tasks.^2,40–44^ Interestingly, even without measures of acoustics/prosody such as speech pauses, articulation rate, and syllable duration (that are technically difficult to code and quantify), we were able to differentiate between nfvPPA, PSP, and CBS versus svPPA, lvPPA, and controls using a simple quantification of connected speech (e.g., type/token count).

Despite a sparse literature on connected speech in PSP and CBS, reduced language output and speech rate have been reported in both groups.^12,17,19^ In the present study, PSP and CBS patients were comparable to nfvPPA patients in that all groups produced fewer words with reduced speech complexity. Our results support previous findings^19,45^ that a general reduction in language output may be a characteristic pattern of PSP and CBS patients, like those with nfvPPA. Moreover, overall performance on various cognitive and language assessments has also been reported to be similar for PSP, CBS and nfvPPA patients.^11,20,46^

### Lexico-semantic features

SvPPA and lvPPA patients produced a greater proportion of words that are more frequent and semantically diverse, as well as shorter and less phonologically complex. This finding is consistent with previous reports and highlights two important points.^4,7^ First, the secondary changes in other psycholinguistic properties such as imageability and length may be related to the under-sampling of the low frequency words used by controls; in other words, svPPA patients generated more “lighter” words that tend to be less imageable and more semantically diverse (e.g., “something”). In addition to under-sampling the low frequency space, svPPA patients have also been found to over-sample the higher frequency space by substituting alternatives to the low frequency target items or picture elements they are unable to name.^4^ For example, in the present study, svPPA patients tended to replace low frequency words typically produced by controls (e.g., “the *sink* is *overflowing*”) with higher frequency words that are less imageable and shorter (e.g., “*it*’s *coming* out”).^47^ Additionally, prior studies have consistently reported that patients with svPPA/semantic dementia replace content words with high frequency, high semantic diversity, and low imageability words not only during picture description, but also in other aspects of language output such as naming and verbal fluency.^4,5,8,39,48,49^ Frequency and age of acquisition effects in svPPA have also been found beyond tests requiring language output such as lexical decision.^50^ Less is known about the psycholinguistic properties of words used by patients with lvPPA. Our findings accord with those of Cho *et al*.^51^ who reported that lvPPA patients produced shorter and more frequent content words when describing the ‘cookie theft’ picture. Furthermore, our formal distribution analysis with the difference plots (Figure 2) and quantification of words produced in each quartile (Figure 3) revealed contrastive patterns across the patient groups with (1) svPPA and lvPPA producing shorter words with high frequency and semantic diversity and (ii) nfvPPA, PSP, and CBS producing later acquired, lower frequency, and less semantically diverse words.

### Grey matter correlates of connected speech features

High scores on the ‘speech quanta’ PC correlated with greater grey matter intensities of bilateral middle and superior frontal gyri, and right inferior frontal gyrus (IFG) extending medially and subcortically to include the insula. Cho *et al*.^40^ found increased speech errors and production of partial words in nfvPPA to be associated with cortical thinning in the left middle frontal gyrus. Ash *et al*.^2^ found speech sound deficits and reduced speech rate in nfvPPA to be related to atrophy in the insula, a region thought to be important for speech articulation,^52,53^ and right premotor and supplementary motor regions. Prior studies have also suggested the role of the superior and middle frontal gyri in the grammatical processing of language production and comprehension.^54,55^ These findings highlight the potential role of the bilateral frontal region in measures of speech production and rate.

High scores on the ‘speech complexity’ PC correlated with grey matter intensities of the left insula, IFG, superior temporal gyrus (STG), and limbic structures. The largest cluster was found for the left insula and IFG, extending into the temporal lobe. Beyond overt speech production, the IFG and insula are reported to be critical in the acoustic measures of speech production such as pause segment duration in motor speech disorders including nfvPPA, ALS, and post-stroke aphasia.^52,56,57^ Our findings are in line with previously reported associations between superior temporal regions and greater morpho-syntactic demands,^58^ grammaticality,^2^ complex sentence production,^59^ lexical phonology^60^, and verbal generation in controls and diverse patient groups.^45^ The STG has also been reported to be implicated in the prefrontal-temporal feedback loop and associated with self-monitoring of speech ouput.^61^

High scores on the ‘length’ PC correlated with greater grey matter intensities of the bilateral temporal lobe, including medial temporal regions, insula, and right limbic lobe. Notably, when excluding controls, the only cluster that correlated significantly included the left insula, middle and superior temporal gyri (see Supplementary Table 6). During an overt picture naming task, Wilson *et al*.^62^ found word length to be positively correlated with signal intensity in the left STG in healthy controls. In addition, Hodgson *et al*.^63^ found the middle and superior temporal regions to be not only implicated in phonology but also general semantics and semantic control. The ability to generate longer, phonologically more complex words and word combinations may rely on processing speech sounds, as well as accessing conceptual knowledge and controlled retrieval of meaningful semantic information.

### Word checklist for picture description

Validated tools to analyse connected speech samples are scare, and to this end, we optimised simple checklists for two widely used picture-narratives to assess PPA subtypes, PSP, and CBS. We employed a hierarchical structure in our LASSO analysis given the nature of word usage across patient groups. The LASSO models could not differentiate svPPA versus lvPPA, nfvPPA versus PSP, and nfvPPA versus PSP and CBS with the target words alone. Supplementing the checklist with MLSE and ACE-R subtest scores improved the differentiation between these groups with excellent within group four-fold cross validation accuracies. Out-of-sample test accuracy was also found to be high for svPPA versus lvPPA, which emphasises the need for further specialist assessments for aphasic groups that cluster based on shared clinical features (i.e., anomia in svPPA and lvPPA, motor speech and/or agrammatism in nfvPPA, PSP, and CBS).

Clinical tools that are fast, simple, and sensitive to aphasia subtypes including various checklists have previously been proposed for post-stroke aphasia,^64^ but to our knowledge this is the first study to provide a direct comparison of word usage across PPA subtypes and Parkinson-plus disorders and optimise a checklist for these patient groups. Future studies with connected speech samples could employ similar methodologies such as our LASSO models to generate specific word checklists for other picture description tasks, different languages, and/or diverse patient groups. The present study could also potentially inform the design of future studies in developing targeted pictures that contain the key vocabulary items that help to differentiate specific clinical groups.

### Limitations and clinical implications

There are limitations to our study. We only present clinical, not pathological, diagnoses, although clinic-pathological correlations are high for PPA and PSP. Our sample size for the out-of-sample test validation was small particularly for certain groups such as PSP. However, we mitigated the potential limitations of small-sample *k*-fold cross-validation by conducting predictive validity testing on an unseen dataset. This supports generalisability of our models and word checklists. Future work is warranted to test the generalisability of the word checklists to larger patient samples that span various disease stages, non-English languages, and varying levels of demographics including education and geographical regions.

A major aim of the present study was to ameliorate the problem of connected speech analyses being time-consuming, effortful and inconsistent across clinicians and different clinical/research settings. As a result, our systematic analysis of connected speech did not include other acoustic and articulatory measures investigated in prior studies. There are undoubtedly other features of the patients’ connected speech that can help with differentiation^43^ which are not captured in our approach, but these require expertise and time-consuming transcription and analyses. Finally, we acknowledge that our imaging analyses were exploratory but nonetheless add to the current literature pertaining to regions engaged in connected speech.

In conclusion, we propose that screening for language deficits in PPA and “motor” disorders like PSP and CBS is achievable with a one-minute sample of connected speech. By focusing on the number and lexico-semantic metrics of the given words, rather than acoustic features, this method is likely to be robust to detect dysarthrophonia from disease, even with reduced bandwidth from remote recordings. The screening test is not a substitute for in-depth neuropsychological assessment, but a contributing tool towards diagnosis. Furthermore, it has the advantage of applicability in resource-limited settings and with limited expertise. Future versions of the test for non-English speakers would further increase the international utility of this approach.

## Data availability

Unthresholded statistical parametric images are available freely on request of the corresponding or senior author. Word lists produced by participants are available on request. Participant MRI scans and transcripts may be available, subject to a data sharing agreement required to protect confidentiality and adhere to consent terms.

## Acknowledgements

We thank our patients and their families for supporting this work.

## Supplementary material

**Supplementary Table 1.** Loadings for PCA of quantitative measures of speech fluency.

| Measure | PC 1 ("Speech Quanta") | PC 2 ("Lexical Richness") | PC 3 ("Speech Complexity") |
| --- | --- | --- | --- |
| Number of Words | **0.97** | -0.21 | 0.00 |
| Number of Word Bigrams | **0.97** | -0.21 | 0.00 |
| Number of Word Trigrams | **0.97** | -0.20 | 0.00 |
| Type of Words | **0.97** | 0.00 | 0.00 |
| Type of Word Bigrams | **0.98** | -0.14 | 0.00 |
| Type of Word Trigrams | **0.98** | -0.16 | 0.00 |
| Combination Ratio | **0.66** | 0.00 | **0.52** |
| Word Per Minute | **0.60** | 0.00 | **0.72** |
| Total Time | 0.48 | -0.11 | **-0.81** |
| TTR of Words | **-0.67** | **0.59** | 0.00 |
| TTR of Word Bigrams | -0.32 | **0.91** | 0.00 |
| TTR of Word Trigrams | 0.00 | **0.93** | 0.00 |
| Proportion of Function Words | 0.40 | -0.19 | 0.29 |

Rotation: Orthogonal varimax. Loadings above a threshold of 0.5 are bolded. PC, principal component; TTR, type-to-token ratio.

**Supplementary Table 2.** Correlations between MLSE and PC scores.

|  | MLSE Motor<br>speech | MLSE Syntax | MLSE Semantics | MLSE Phonology | MLSE Working<br>Memory | MLSE Total |
| --- | --- | --- | --- | --- | --- | --- |
| **Speech fluency PCA** |  |  |  |  |  |  |
| PC 1 'speech quanta' |  |  |  |  |  |  |
| All groups | **R = 0.54,**<br>**P < 0.001** | **R = 0.42,**<br>**P < 0.001** | R = 0.11,<br>P = 0.38 | **R = 0.5,**<br>**P < 0.001** | R = 0.08,<br>P = 0.51 | **R = 0.45,**<br>**P < 0.001** |
| Controls | R = -0.08, P = 0.71 | R = 0.29,<br>P = 0.16 | R = -0.15,<br>P = 0.47 | R = 0.23,<br>P = 0.28 | R = -0.16,<br>P = 0.45 | R = 0.09,<br>P = 0.69 |
| svPPA | R = 0.23,<br>P = 0.59 | R = -0.21, P = 0.61 | R = 0.23,<br>P = 0.59 | R = 0.38,<br>P = 0.36 | R = -0.65,<br>P = 0.08 | R = -0.03,<br>P = 0.94 |
| lvPPA | **R = 0.83,**<br>**P = 0.02** | R = 0.44,<br>P = 0.33 | R = 0.62,<br>P = 0.14 | **R = 0.94,**<br>**P = 0.002** | R = -0.23,<br>P = 0.62 | **R = 0.88,**<br>**P = 0.009** |
| nvPPA | R = -0.27, P = 0.52 | R = -0.01, P = 0.99 | R = 0.41,<br>P = 0.32 | R = -0.3,<br>P = 0.48 | R = -0.16,<br>P = 0.71 | R = -0.19,<br>P = 0.65 |
| PSP | R = 0.12,<br>P = 0.75 | R = -0.17, P = 0.65 | R = 0.44,<br>P = 0.24 | R = 0.3,<br>P = 0.44 | R = -0.02,<br>P = 0.96 | R = 0.09,<br>P = 0.81 |
| CBS | R = 0.49,<br>P = 0.15 | R = 0.31,<br>P = 0.38 | R = -0.35,<br>P = 0.32 | R = 0.16,<br>P = 0.65 | R = -0.51,<br>P = 0.13 | R = 0.13,<br>P = 0.71 |
| PC 2 'lexical richness' |  |  |  |  |  |  |
| All groups | R = -0.16, P = 0.21 | R = 0.05,<br>P = 0.68 | **R = 0.44,**<br>**P < 0.001** | R = 0.03,<br>P = 0.8 | R = -0.01,<br>P = 0.92 | R = 0.14,<br>P = 0.27 |
| Controls | R = 0.13,<br>P = 0.53 | R = -0.11, P = 0.6 | R = -0.07,<br>P = 0.73 | R = -0.35,<br>P = 0.10 | R = -0.37,<br>P = 0.07 | R = -0.36,<br>P = 0.09 |
| svPPA | R = 0.02,<br>P = 0.97 | R = 0.15,<br>P = 0.73 | R = -0.31,<br>P = 0.46 | R = 0.06,<br>P = 0.89 | R = -0.17,<br>P = 0.69 | R = -0.19,<br>P = 0.66 |
| lvPPA | R = 0.66,<br>P = 0.1 | R = 0.45,<br>P = 0.31 | R = 0.73,<br>P = 0.06 | **R = 0.97,**<br>**P < 0.001** | R = -0.3,<br>P = 0.52 | **R = 0.96,**<br>**P < 0.001** |
| nvPPA | R = -0.58, P = 0.13 | R = -0.28, P = 0.5 | R = 0.38,<br>P = 0.34 | R = -0.37,<br>P = 0.36 | R = -0.5,<br>P = 0.21 | R = -0.44,<br>P = 0.27 |
| PSP | R = 0.34,<br>P = 0.37 | R = 0.22,<br>P = 0.56 | R = 0.12,<br>P = 0.75 | R = 0.47,<br>P = 0.21 | R = 0.11,<br>P = 0.77 | R = 0.4,<br>P = 0.29 |
| CBS | R = -0.29, P = 0.42 | R = 0.16,<br>P = 0.67 | R = 0.25,<br>P = 0.48 | R = 0.12,<br>P = 0.74 | R = 0.52,<br>P = 0.12 | R = 0.09,<br>P = 0.81 |
| PC 3 'speech complexity' |  |  |  |  |  |  |
| All groups | **R = 0.49,**<br>**P < 0.001** | **R = 0.52,**<br>**P < 0.001** | **R = 0.29,**<br>**P = 0.02** | **R = 0.53,**<br>**P < 0.001** | **R = 0.5,**<br>**P < 0.001** | **R = 0.62,**<br>**P < 0.001** |
| Controls | R = 0.34,<br>P = 0.1 | R = -0.03, P = 0.89 | R = -0.09,<br>P = 0.67 | R = 0.1,<br>P = 0.63 | **R = 0.49,**<br>**P = 0.01** | R = 0.28,<br>P = 0.18 |
| svPPA | R = -0.35, P = 0.4 | R = -0.18, P = 0.66 | R = -0.17,<br>P = 0.7 | R = 0.14,<br>P = 0.74 | R = 0.04,<br>P = 0.93 | R = -0.12,<br>P = 0.79 |
| lvPPA | R = -0.02, P = 0.97 | R = -0.33, P = 0.47 | R = -0.25,<br>P = 0.59 | R = -0.53,<br>P = 0.22 | R = -0.02,<br>P = 0.97 | R = -0.41,<br>P = 0.36 |
| nvPPA | R = 0.36,<br>P = 0.38 | R = 0.69,<br>P = 0.06 | R = 0.62,<br>P = 0.10 | **R = 0.9,**<br>**P = 0.003** | R = 0.63,<br>P = 0.09 | **R = 0.77,**<br>**P = 0.03** |
| PSP | **R = 0.88,**<br>**P = 0.002** | R = 0.34,<br>P = 0.37 | R = 0.37,<br>P = 0.33 | R = 0.34,<br>P = 0.36 | R = 0.3,<br>P = 0.43 | **R = 0.75,**<br>**P = 0.02** |
| CBS | **R = 0.66,**<br>**P = 0.04** | R = 0.44,<br>P = 0.21 | **R = 0.83,**<br>**P = 0.003** | **R = 0.88,**<br>**P < 0.001** | R = 0.51,<br>P = 0.13 | **R = 0.91,**<br>**P < 0.001** |
| **Word Properties PCA** |  |  |  |  |  |  |
| PC 1 'length' |  |  |  |  |  |  |
| All groups | R = 0.08,<br>P = 0.53 | R = 0.14,<br>P = 0.26 | **R = 0.56,**<br>**P < 0.001** | **R = 0.24,**<br>**P = 0.05** | **R = 0.26,**<br>**P = 0.03** | **R = 0.38,**<br>**P = 0.001** |
| Controls | R = -0.07, P = 0.74 | R = -0.26, P = 0.21 | R = -0.16,<br>P = 0.46 | R = 0.02,<br>P = 0.94 | R = -0.21,<br>P = 0.32 | R = -0.17,<br>P = 0.42 |
| svPPA | R = -0.29, P = 0.49 | R = -0.38, P = 0.35 | R = -0.04,<br>P = 0.92 | R = -0.64,<br>P = 0.09 | R = 0.18,<br>P = 0.68 | R = -0.33,<br>P = 0.43 |
| lvPPA | R = 0.38,<br>P = 0.36 | R = -0.34, P = 0.41 | R = 0.62,<br>P = 0.10 | R = 0.6,<br>P = 0.12 | R = -0.06,<br>P = 0.89 | R = 0.59,<br>P = 0.13 |
| nvPPA | R = -0.05, P = 0.9 | R = 0.11,<br>P = 0.79 | R = -0.32,<br>P = 0.41 | R = 0.07,<br>P = 0.85 | R = 0.13,<br>P = 0.74 | R = 0.04,<br>P = 0.91 |
| PSP | R = -0.4,<br>P = 0.29 | R = -0.24, P = 0.54 | R = -0.26,<br>P = 0.49 | R = -0.09,<br>P = 0.81 | R = -0.36,<br>P = 0.34 | R = -0.34,<br>P = 0.38 |
| CBS | R = 0.43,<br>P = 0.21 | R = -0.04, P = 0.91 | **R = 0.63,**<br>**P = 0.05** | R = 0.58,<br>P = 0.08 | R = 0.3,<br>P = 0.4 | R = 0.57,<br>P = 0.09 |
| PC 2 'semantic richness' |  |  |  |  |  |  |
| All groups | **R = 0.26,**<br>**P = 0.03** | R = -0.08, P = 0.52 | **R = -0.54,**<br>**P < 0.001** | R = 0.17,<br>P = 0.16 | R = -0.21,<br>P = 0.09 | R = -0.15,<br>P = 0.23 |
| Controls | R = -0.06, P = 0.78 | R = 0.16,<br>P = 0.45 | R = 0.26,<br>P = 0.21 | R = 0.1,<br>P = 0.63 | R = -0.31,<br>P = 0.15 | R = -0.002, P = 0.99 |
| svPPA | R = -0.10, P = 0.82 | R = -0.64, P = 0.09 | R = -0.33, P = 0.42 | R = 0.33, P = 0.43 | **R = -0.76, P = 0.03** | R = -0.58, P = 0.13 |
| lvPPA | R = 0.65, P = 0.08 | R = -0.10, P = 0.82 | R = -0.03, P = 0.94 | R = 0.62, P = 0.1 | R = 0.11, P = 0.79 | R = 0.28, P = 0.5 |
| nvPPA | R = -0.09, P = 0.82 | R = 0.07, P = 0.86 | R = 0.03, P = 0.93 | R = 0.3, P = 0.43 | R = -0.007, P = 0.98 | R = 0.09, P = 0.81 |
| PSP | R = 0.03, P = 0.94 | R = -0.63, P = 0.07 | R = 0.07, P = 0.86 | R = -0.07, P = 0.85 | R = -0.23, P = 0.55 | R = -0.16, P = 0.68 |
| CBS | R = 0.06, P = 0.88 | R = 0.59, P = 0.08 | R = -0.13, P = 0.73 | R = -0.004, P = 0.99 | R = 0.02, P = 0.96 | R = 0.08, P = 0.83 |
| PC 3 'acquisition age' |  |  |  |  |  |  |
| All groups | **R = 0.55, P < 0.001** | R = 0.23, P = 0.06 | R = -0.09, P = 0.46 | **R = 0.53, P < 0.001** | R = 0.09, P = 0.46 | **R = 0.33, P = 0.007** |
| Controls | R = 0.07, P = 0.74 | R = 0.34, P = 0.1 | R = 0.02, P = 0.94 | R = -0.11, P = 0.59 | R = -0.34, P = 0.1 | R = -0.12, P = 0.59 |
| svPPA | R = 0.08, P = 0.85 | R = -0.2, P = 0.64 | R = -0.22, P = 0.61 | R = 0.42, P = 0.3 | R = -0.14, P = 0.74 | R = -0.09, P = 0.84 |
| lvPPA | **R = 0.84, P = 0.009** | R = 0.11, P = 0.8 | R = 0.4, P = 0.32 | **R = 0.86, P = 0.006** | R = -0.16, P = 0.7 | R = 0.6, P = 0.12 |
| nvPPA | R = 0.37, P = 0.32 | R = -0.29, P = 0.44 | R = -0.12, P = 0.75 | R = 0.28, P = 0.47 | R = 0.3, P = 0.44 | R = 0.23, P = 0.55 |
| PSP | R = -0.22, P = 0.57 | R = -0.31, P = 0.41 | R = -0.02, P = 0.97 | R = 0.45, P = 0.23 | R = 0.27, P = 0.48 | R = -0.008, P = 0.98 |
| CBS | R = 0.57, P = 0.08 | **R = 0.71, P = 0.02** | R = 0.41, P = 0.24 | R = 0.55, P = 0.1 | R = 0.16, P = 0.66 | **R = 0.65, P = 0.04** |

Note: Significant correlations are indicated in bold font.

**Supplementary Table 3.** Loadings for PCA of quantitative measures of word properties.

| Measure | PC 1 ("Length") | PC 2 ("Semantic richness") | PC 3 ("Acquisition age") |
| --- | --- | --- | --- |
| Length | **0.89** | -0.14 | 0.22 |
| OLD | **0.95** | 0.00 | 0.19 |
| PLD | **0.94** | 0.00 | 0.16 |
| Log Frequency | -0.28 | **0.88** | -0.22 |
| Semantic Diversity | 0.00 | **0.86** | 0.23 |
| SND | -0.19 | **0.84** | -0.30 |
| Concreteness | -0.18 | **-0.65** | **-0.59** |
| Age of Acquisition | 0.35 | -0.17 | **0.81** |

Rotation: Orthogonal varimax. Loadings above a threshold of 0.5 are bolded. OLD, orthographic Levenshtein distance; PC, principal component; PLD, phonological Levenshtein distance; SND, semantic neighbourhood density.

**Supplementary Table 4.** Distribution analysis. ANOVA findings on the effects of group, quartile and group-by-quartile interaction from the distribution analysis of word properties principal component analysis

| Principal Component (PC) | Task | ANOVA | Tukey's HSD Test for multiple comparison |
| --- | --- | --- | --- |
| **PC 1 ('Length')** | BDAE 'cookie theft' | Effect of group only: ($F(5,283) = 37.16, p < 0.001$) | Controls > all patients ($p < 0.001$), svPPA > nfvPPA, PSP and CBS ($p < 0.01$), lvPPA > nfvPPA ($p = 0.005$) |
| | MLSE 'beach scene' | Effect of group only: ($F(5,272) = 39.18, p < 0.001$) | Controls > all patients ($p < 0.001$), svPPA > nfvPPA, PSP and CBS ($p \leq 0.001$), lvPPA and CBS > nfvPPA ($p < 0.05$) |
| **PC 2 ('Semantic richness')** | BDAE 'cookie theft' | Effects of group ($F(5,280) = 33.68, p < 0.001$), quartile ($F(1,280) = 4.67, p = 0.03$), and group-by-quartile interaction ($F(5,280) = 4.36, p < 0.001$) | For group: Controls > all patients ($p < 0.001$), svPPA > nfvPPA, PSP and CBS ($p < 0.005$), lvPPA > nfvPPA ($p < 0.02$)<br>For quartile: first > second ($p = 0.05$), third > second ($p = 0.02$), fourth > second ($p < 0.001$) |
| | MLSE 'beach scene' | Effects of group ($F(5,270) = 28.94, p < 0.001$), quartile ($F(1,270) = 5.53, p = 0.02$), and group-by-quartile interaction ($F(5,270) = 8.29, p < 0.001$). | For group: Controls > all patients ($p \leq 0.005$), svPPA > nfvPPA, PSP and CBS ($p < 0.01$), lvPPA > nfvPPA ($p < 0.001$)<br>For quartile: second > first ($p = 0.007$), second > third ($p = 0.007$), second > fourth ($p < 0.001$) |
| **PC 3 ('Acquisition Age')** | BDAE 'cookie theft' | Effects of group ($F(5,283) = 36.15, p < 0.001$), quartile ($F(1,283) = 17.17, p < 0.001$), and group-by-quartile interaction ($F(5,283) = 2.47, p = 0.03$) | For group: Controls > all patients ($p < 0.001$), svPPA > nfvPPA, PSP and CBS ($p < 0.01$), lvPPA > nfvPPA ($p < 0.005$)<br>For quartile: third > first ($p = 0.01$), fourth > first ($p = 0.01$), third > second ($p = 0.002$), fourth > second ($p = 0.002$) |
| | MLSE 'beach scene' | Effects of group ($F(5,265) = 31.04, p < 0.001$), quartile ($F(1,265) = 21.67, p < 0.001$), and group-by-quartile interaction ($F(5,265) = 2.47, p = 0.03$) | For group: Controls > all patients ($p \leq 0.007$), svPPA > nfvPPA, PSP and CBS ($p < 0.01$), lvPPA > nfvPPA ($p < 0.001$)<br>For quartile: first > third ($p = 0.01$), first > fourth ($p < 0.001$), second > third ($p < 0.001$), second > fourth ($p < 0.001$) |

**Supplementary Table 5.** VBM results in the whole group. Voxel based morphometry results showing regions of grey matter intensity that correlate with PCA-generated principal component in the whole group

| Principal Component | Regions | Hemisphere | Number of Voxels | Peak MNI coordinates |  |  | t-value |
| --- | --- | --- | --- | --- | --- | --- | --- |
| **Speech quanta (Supplementary Table 2 PC 1)** | Middle and superior frontal gyri | Left | 407 | -22 | 22 | 48 | 5.56 |
|  | Middle and superior frontal gyri and supplementary motor area | Right | 287 | 18 | 26 | 58 | 4.25 |
|  | Inferior frontal gyrus and insula | Right | 235 | 36 | 24 | 6 | 4.95 |
|  | Putamen and caudate | Right | 229 | 20 | 14 | 0 | 5.54 |
| **Speech complexity (Supplementary Table 2 PC 3)** | Insula, inferior frontal gyrus, extending into the superior temporal gyrus | Left | 1405 | -44 | 6 | 4 | 5.65 |
|  | Medial frontal gyrus, superior frontal gyrus, and anterior cingulate | Left | 245 | -4 | 54 | -2 | 4.97 |
|  | Middle and superior frontal gyri | Left | 115 | -24 | 34 | 44 | 4.21 |
|  | Parahippocampal gyrus, amygdala and hippocampus | Left | 109 | -26 | -10 | -12 | 4.24 |
| **Length (Supplementary Table 3 PC 1)** | Insula, middle and superior temporal gyri | Left | 828 | -44 | -6 | -8 | 5.32 |
|  | Parahippocampal and fusiform gyri | Left | 356 | -24 | -34 | -20 | 4.99 |
|  | Limbic lobe, including the anterior cingulate and caudate | Right | 236 | 4 | 12 | -10 | 4.45 |
|  | Inferior and middle temporal gyri and fusiform gyri | Right | 200 | 46 | -10 | -38 | 4.52 |
|  | Parahippocampal gyrus, hippocampus, fusiform and amygdala | Right | 139 | 30 | -12 | -32 | 4.05 |
| **Acquisition age (Supplementary Table 3 PC 3)** | Cingulate gyrus | Bilateral | 196 | 2 | -8 | 44 | 4.35 |
|  | Caudate and putamen | Right | 102 | 16 | 14 | 6 | 4.12 |

**Supplementary Table 6.** VBM results in patients. Voxel based morphometry results showing regions of grey matter intensity that correlate with PCA-generated factors in patients only

| Principal Component | Regions | Hemisphere | Number of Voxels | Peak MNI coordinates |  |  | t-value |
| --- | --- | --- | --- | --- | --- | --- | --- |
| **Length**<br>**(Supplementary Table 3 PC 1)** | Insula, middle and superior temporal gyri | Left | 184 | -46 | -8 | -6 | 4.69 |

**Supplementary Table 7.** VBM results with cluster-forming height threshold. Voxel based morphometry results showing regions of grey matter intensity that correlate with PCA-generated factors with a cluster-forming height threshold of *p* < 0.005 paired with a cluster extent threshold of *p* < 0.05 FWE-corrected

| Principal Component | Regions | Hemisphere | Number of Voxels | Peak MNI coordinates |  |  | t-value |
| --- | --- | --- | --- | --- | --- | --- | --- |
| **Speech complexity (Supplementary Table 2 PC 3)** | Insula, inferior frontal gyrus, extending into the superior temporal gyrus | Left | 1405 | -44 | 6 | 4 | 5.65 |
| **Length (Supplementary Table 3 PC 1)** | Insula, middle and superior temporal gyri | Left | 828 | -44 | -6 | -8 | 5.32 |

**Supplementary Table 8.** LASSO results comparing all patients versus controls, “lexico-semantic” (svPPA and lvPPA) versus “motor” (nfvPPA, PSP, and CBS) groups, svPPA versus lvPPA patients, and nfvPPA and PSP versus CBS patients.

|  | Word checklist:<br>LASSO value | Word checklist with cognitive<br>scores: LASSO value |
| --- | --- | --- |
| **I. BDAE ‘cookie theft’** |  |  |
| **Controls versus patients** |  |  |
| Model intercept | -5.99 | -7.76 |
| Overflow | 2.94 | 0.26 |
| Stool | 1.53 |  |
| Open | 1.92 | 0.44 |
| Not | 0.25 |  |
| Water | 0.14 |  |
| Sink | 0.39 | 0.27 |
| Doing | 0.81 | 0.31 |
| Something | -0.60 |  |
| Drying | 1.91 | 1.92 |
| Over | 0.33 | 0.02 |
| Garden | 0.45 | 0.37 |
| MLSE: Syntax |  | 0.01 |
| MLSE: Working memory |  | 0.07 |
| ACE-R: Fluency |  | 0.55 |
| **“Motor” (nfvPPA, PSP, CBS) versus “Lexico-semantic” (svPPA, lvPPA)** |  |  |
| Model intercept | -2.07 | 1.91 |
| Overflow | -1.47 |  |
| Stool | -1.11 | -0.23 |
| Open | -0.53 |  |
| Not | 1.92 | 1.09 |
| Water | 0.72 |  |
| Sink | -0.40 |  |
| Something | 0.17 |  |
| Has/have | 1.64 | 0.69 |
| Little | 1.07 | 0.91 |
| Lady | 1.40 |  |
| MLSE: Motor speech |  | 0.02 |
| MLSE: Semantics |  | -0.27 |
| **svPPA versus lvPPA** |  |  |
| Model intercept | NA | -1.56 |
| Drying |  | 1.12 |
| MLSE: Semantics |  | -0.21 |
| MLSE: Syntax |  | 0.25 |
| ACE-R: Visuospatial |  | 0.15 |
| **nfvPPA versus PSP and CBS** |  |  |
| Model intercept | NA | -1.12 |
| Stool |  | 0.21 |
| Looking |  | 0.61 |
| Sink |  | -0.33 |
| Doing |  | 0.54 |
| Has/have |  | 0.34 |
| MLSE: Syntax |  | 0.34 |
| **2. MLSE ‘beach scene’** |  |  |
| **Controls versus patients** |  |  |
| Model intercept | -5.28 | -9.04 |
| Sandcastle | 0.37 |  |
| Sun | 1.88 | 1.88 |
| Dig | 1.12 |  |
| Book | 0.37 |  |
| Sand | 0.15 |  |
| Sea | 0.59 |  |
| Wearing | 2.30 | 1.34 |
| Bone | 1.26 | 0.49 |
| Towel | 0.32 | 0.56 |
| Beach | 0.75 | 0.86 |
| MLSE: Syntax |  | 0.03 |
| MLSE: Working memory |  | 0.11 |
| ACE-R: Fluency |  | 0.50 |
| **“Motor” (nfvPPA, PSP, CBS) versus “Lexico-semantic” (svPPA, lvPPA)** |  |  |
| Model intercept | -1.31 | 2.50 |
| Sandcastle | -0.16 |  |
| Seagull | -0.96 | -0.30 |
| Do | 1.22 | 0.44 |
| Book | -0.35 | -0.34 |
| Know | 1.31 | 1.64 |
| Beach | 0.40 | 0.38 |
| Got | 1.55 | 2.17 |
| MLSE: Motor speech |  | 0.02 |
| MLSE: Semantics |  | -0.26 |
| ACE-R: Fluency |  | -0.10 |
| ACE-R: Visuospatial |  | -0.04 |
| **svPPA versus lvPPA** |  |  |
| Model intercept | NA | -1.54 |
| Towel |  | -0.20 |
| MLSE: Semantics |  | -0.22 |
| MLSE: Syntax |  | 0.43 |
| ACE-R: Visuospatial |  | 0.08 |
| **nfvPPA and PSP versus CBS** |  |  |
| Model intercept | NA | -0.29 |
| Sandcastle |  | 0.05 |
| Sand |  | 0.47 |
| Towel |  | 0.36 |
| MLSE: Syntax |  | 0.31 |
| ACE-R: Visuospatial |  | -0.05 |

**Supplementary Table 9.**
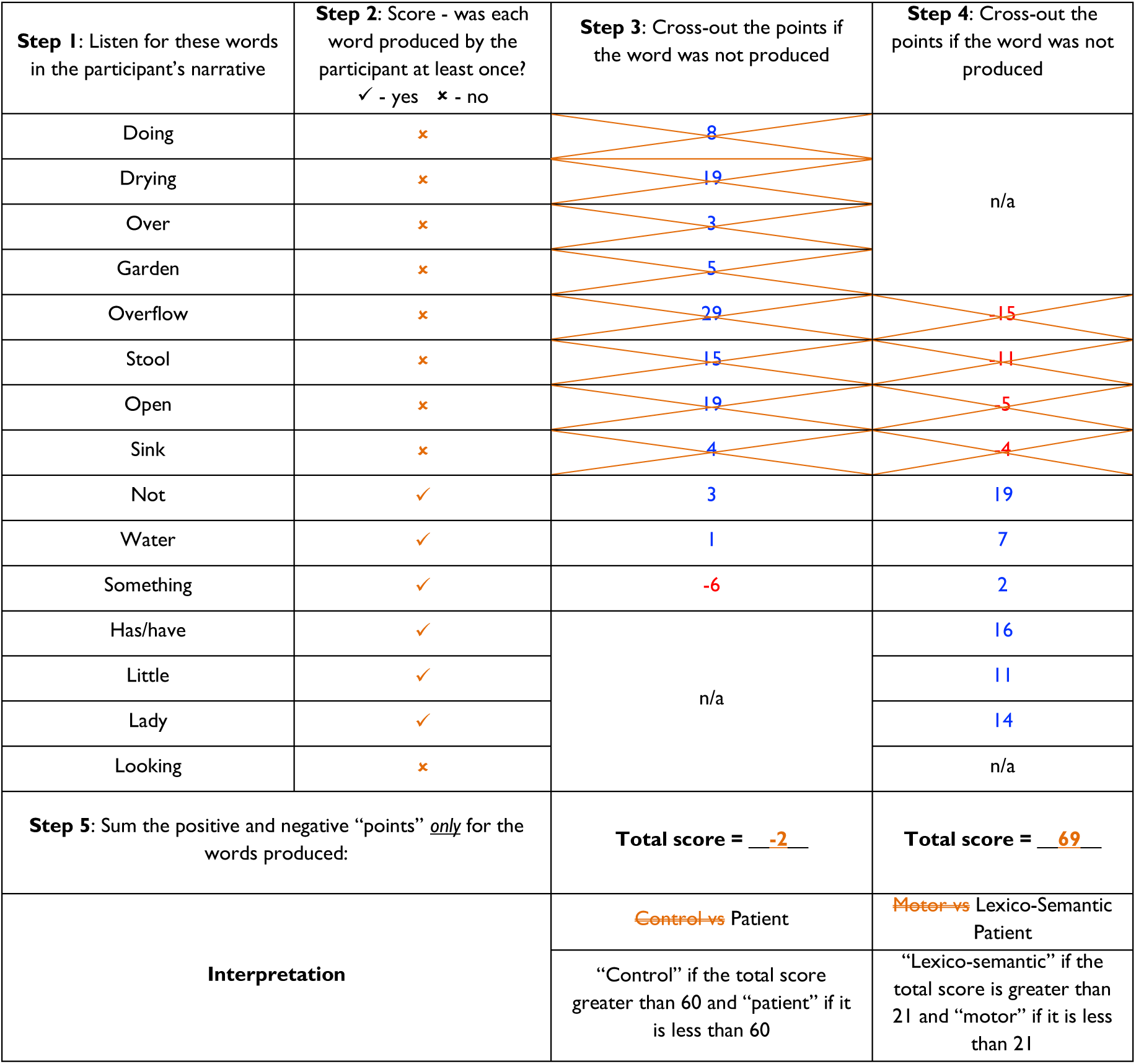
A representative example of an anonymised svPPA patient using the BDAE ‘cookie theft’ 15-word checklist score sheet.

**Supplementary Figure 1.**
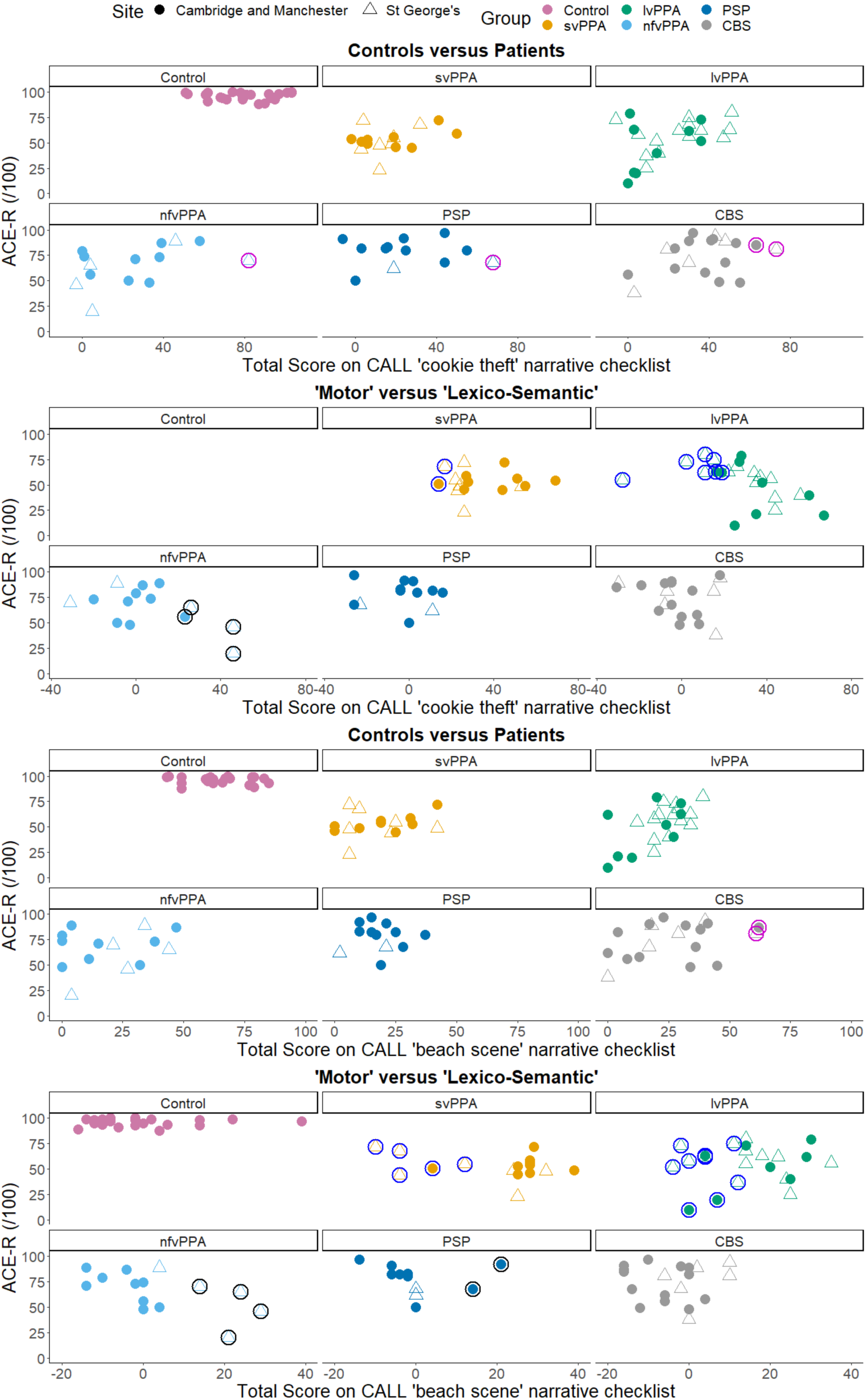
Scatterplots showing total scores on the CALL 15-word checklists and ACE-R with the following color representations: magenta circles for people misclassified as controls, blue circles for those misclassified as belonging to the “motor” group, and black circles for those misclassified as belonging to the “lexico-semantic” group

**Supplementary Figure 2.**
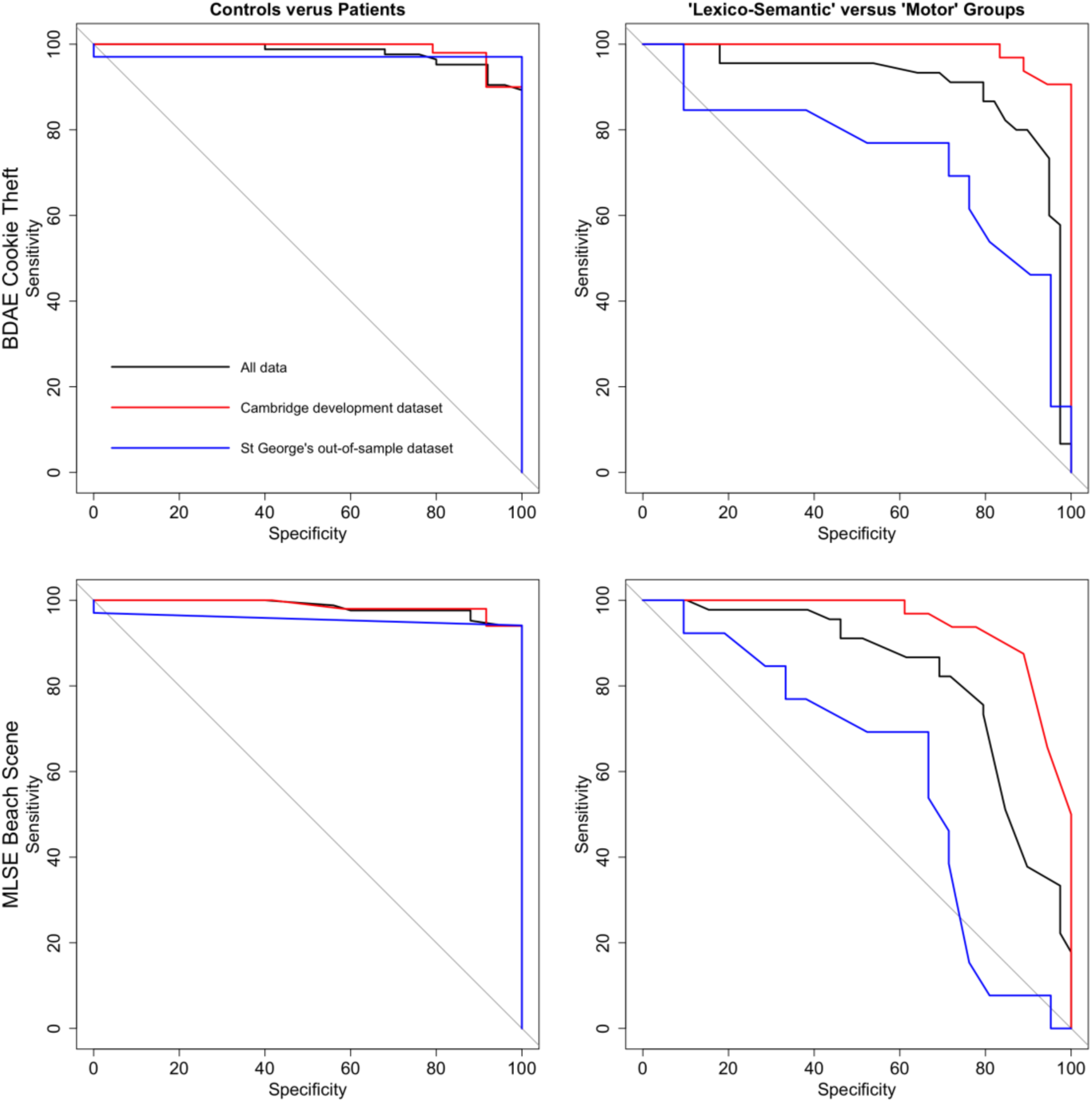
Receiver operating characteristic (ROC) curves showing how well the CALL checklists distinguish between controls and patients (left), and between “lexico-semantic” and “motor” groups (right). All data are shown in black, the Cambridge validation dataset is shown in red, and St George’s out-of-sample dataset is shown in blue. When comparing controls relative to all patients using the BDAE ‘cookie theft’ checklist, the area under the curve (AUC) was largest for the Cambridge development dataset (98.92%), followed by all data (98%) and the St George’s out-of-sample dataset (97.06%). Using the MLSE ‘beach scene’ checklist, the AUC was largest for the Cambridge development dataset (98.67%), followed by all data (98.5%) and the St George’s out-of-sample dataset (95.59%). When comparing “lexico-semantic” versus “motor” groups using the BDAE ‘cookie theft’ checklist, the AUC was largest for the Cambridge development dataset (98.87%), followed by all data (90.23%) and the St George’s out-of-sample dataset (73.63%). Using the MLSE ‘beach scene’ checklist, the AUC was largest for the Cambridge development dataset (94.53%), followed by all data (82.65%) and the St George’s out-of-sample dataset (60.07%).

## Appendix: Cambridge Language List (CALL)

The scoresheets using the 15 words are shown below in Appendices 1 and 2. The first column shows the 15 target words. In the second column, the examiner marks if the participant produced each specific word or not. While variations in morpho-syntactic word forms (e.g., wearing instead of wear/worn) should be marked as correct, synonyms and related words (e.g., “woman” for “lady”) should not. The third column shows the specific “points” associated with each of the words that help to differentiate between healthy controls and patients. The “points” per word were derived directly from the LASSO coefficients. To make CALL easy to use, we rounded up the coefficient and cut-off values to 1 decimal point and then multiplied all values by ten.

As an example, for the BDAE ‘cookie theft’ picture, production of the word “doing” is credited with 8 “points”, whilst the production of “something” is debited with 6 “penalty points”. The total points for the words that each participant produced are calculated. If the summed value exceeds 60 then a “diagnosis” of control is more likely; a score below 60 denotes that the participant is more likely to be a patient. If an indicative diagnosis of patient results (from the scoring of column 3), then a similar secondary scoring process is undertaken. This time, the fourth column provides the positive (blue) and negative (red) “points” associated with the prediction of “lexico-semantic” vs. “motor” patient group membership. Again, the total positive and negative points for the words that each participant produced are calculated. If the summed value exceeds 21 then a “diagnosis” of “lexico-semantic” patient is more likely; a score below 21 denotes that the participant is more likely to be a “motor” patient. For a worked example of a representative svPPA patient, see Supplementary Table 9.

### Appendix 1

Please see below the checklist scoresheets for the (A) BDAE ‘cookie theft’ picture narrative and (B) the MLSE ‘beach scene’ picture narrative differentiating healthy controls versus patients, and between “lexico-semantic” and “motor” groups.

| **(A) Cookie Theft Picture Narrative: Scoresheet for controls versus patients and “lexico-semantic” versus “motor” groups** |  |  |  |
| --- | --- | --- | --- |
| **Step 1:** Listen for these words in the participant’s narrative | **Step 2:** Score - was each word produced by the participant at least once?<br>✓ - yes ✗ - no | **Step 3:** Cross-out the points if the word was not produced | **Step 4:** Cross-out the points if the word was not produced |
| Doing |  | 8 | n/a |
| Drying |  | 19 |  |
| Over |  | 3 |  |
| Garden |  | 5 |  |
| Overflow |  | 29 | -15 |
| Stool |  | 15 | -11 |
| Open |  | 19 | -5 |
| Sink |  | 4 | -4 |
| Not |  | 3 | 19 |
| Water |  | 1 | 7 |
| Something |  | -6 | 2 |
| Has/have |  | n/a | 16 |
| Little |  |  | 11 |
| Lady |  |  | 14 |
| Looking |  |  | n/a |
| **Step 5:** Sum the positive and negative “points” <u>only</u> for the words produced: |  | **Total score = _____** | **Total score = _____** |
| **Interpretation** |  | Control vs Patient | Motor vs Lexico-Semantic Patient |
|  |  | “Control” if the total score is greater than 60 and “patient” if it is less than 60 | “Lexico-semantic” if the total score is greater than 21 and “motor” if it is less than 21 |

### Appendix 2

The checklist scoresheets for additional diagnostic differentiations. As in the main manuscript, we employed a hierarchical classification (i.e., controls versus patients; “lexico-semantic” versus “motor” groups) to the checklists as shown in Appendix 1 as the LASSO regressions for svPPA versus lvPPA, and nfvPPA versus PSP resulted in zero words for both pictures. Under each checklist below, we provide the within-group and out-of-sample validation accuracies to use for reference.

| (B) Beach Scene Picture Narrative: Scoresheet for controls versus patients and “lexico-semantic” versus “motor” groups |  |  |  |
| --- | --- | --- | --- |
| Step 1: Listen for these words in the participant’s narrative | Step 2: Score - was each word produced by the participant at least once?<br>✓ - yes ✗ - no | Step 3: Cross-out the points if the word was not produced | Step 4: Cross-out the points if the word was not produced |
| Seagull |  | n/a | -10 |
| Sandcastle |  | 4 | -2 |
| Book |  | 4 | -4 |
| Sun |  | 19 | n/a |
| Dig |  | 11 |  |
| Bone |  | 13 |  |
| Wearing |  | 23 |  |
| Sand |  | 2 |  |
| Sea |  | 6 |  |
| Towel |  | 3 |  |
| Reading |  | n/a |  |
| Beach |  |  | 4 |
| Know |  |  | 13 |
| Got |  |  | 16 |
| Do |  |  | 12 |
| Step 5: Sum the positive and negative “points” <u>only</u> for the words produced: |  | Total score = _____ | Total score = _____ |
| Interpretation |  | Control vs Patient | Motor vs Lexico-Semantic Patient |
|  |  | “Control” if the total score is greater than 53 and “patient” if it is less than 53 | “Lexico-semantic” if the total score is greater than 13 and “motor” if it is less than 13 |

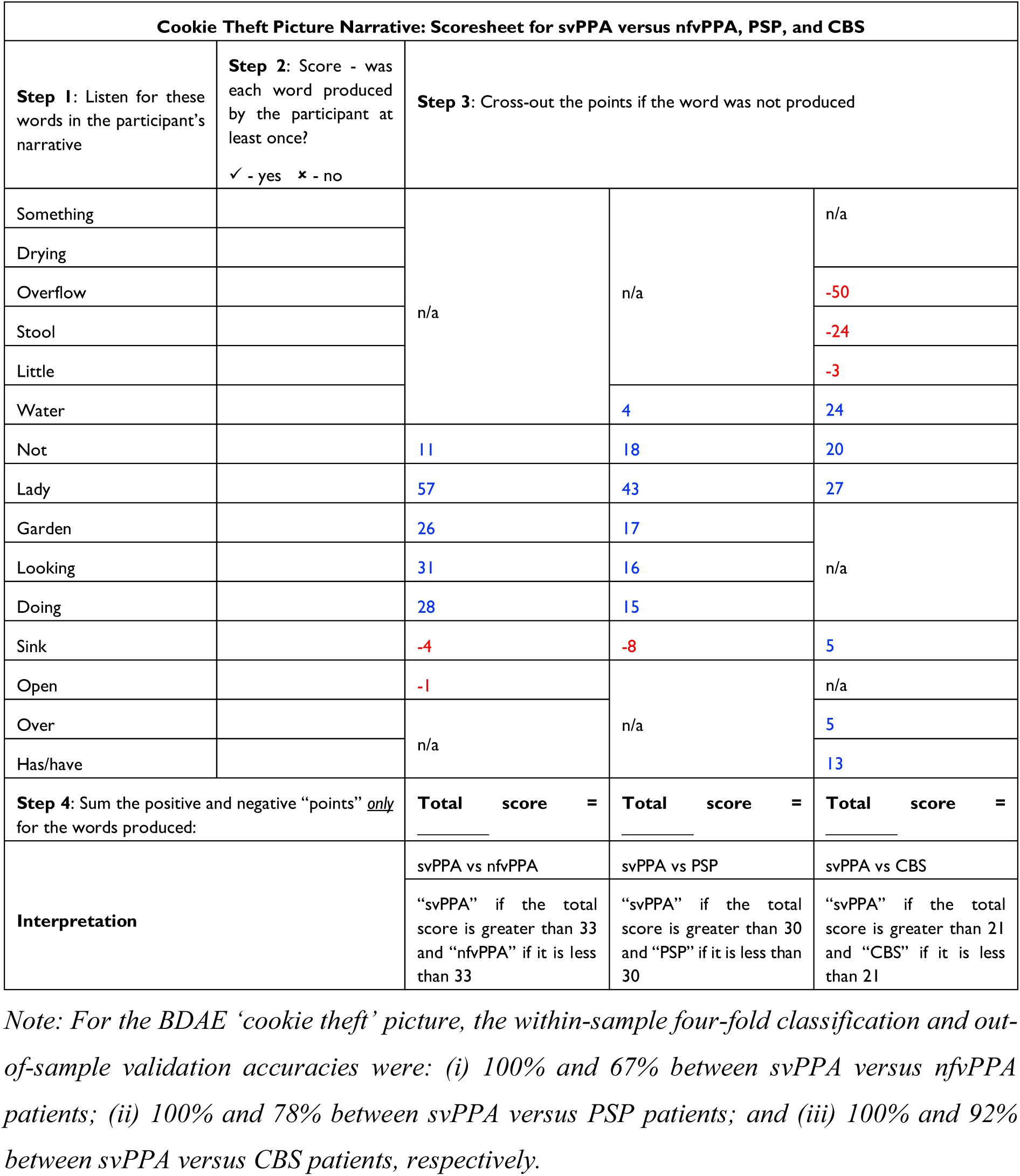

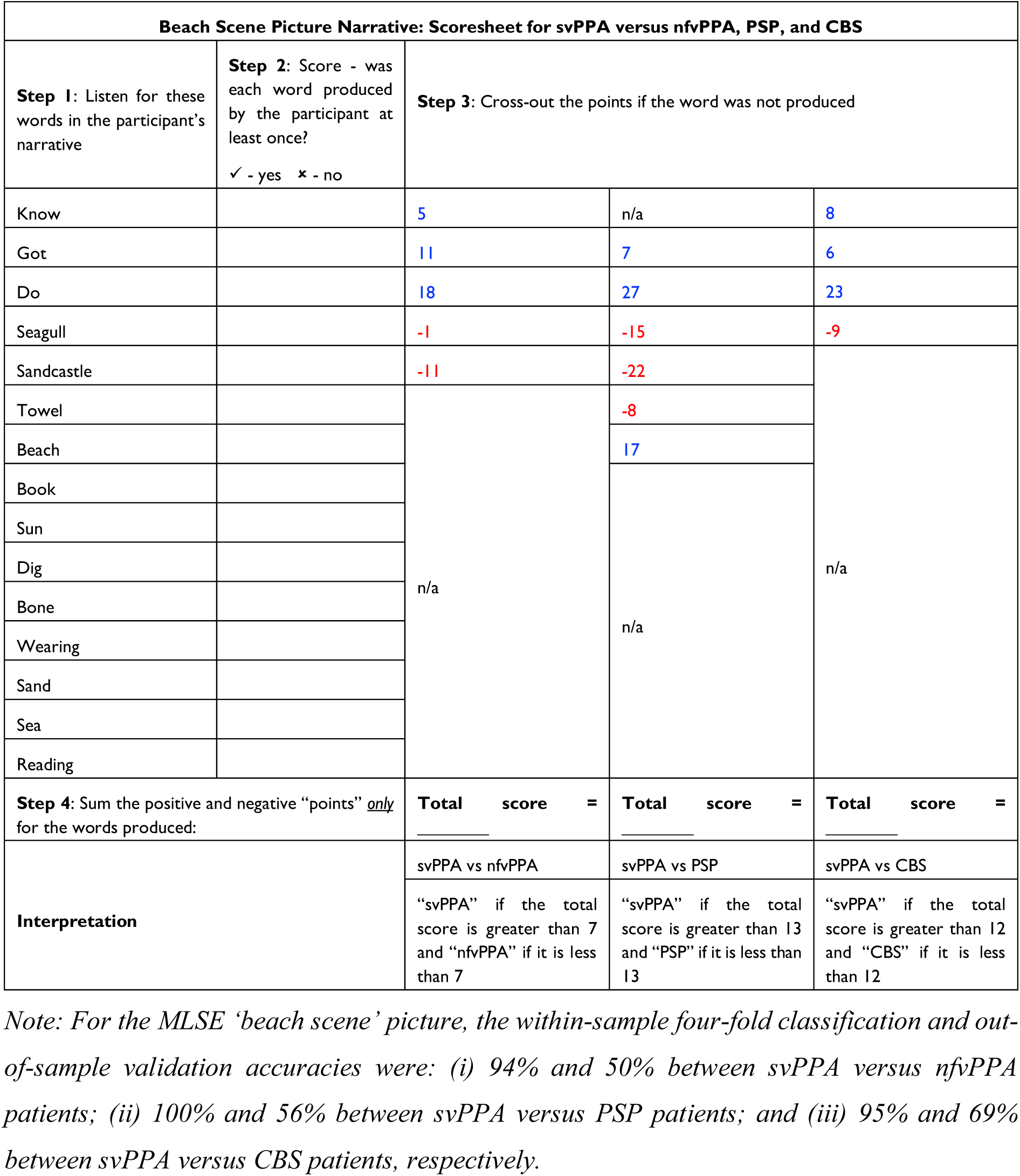

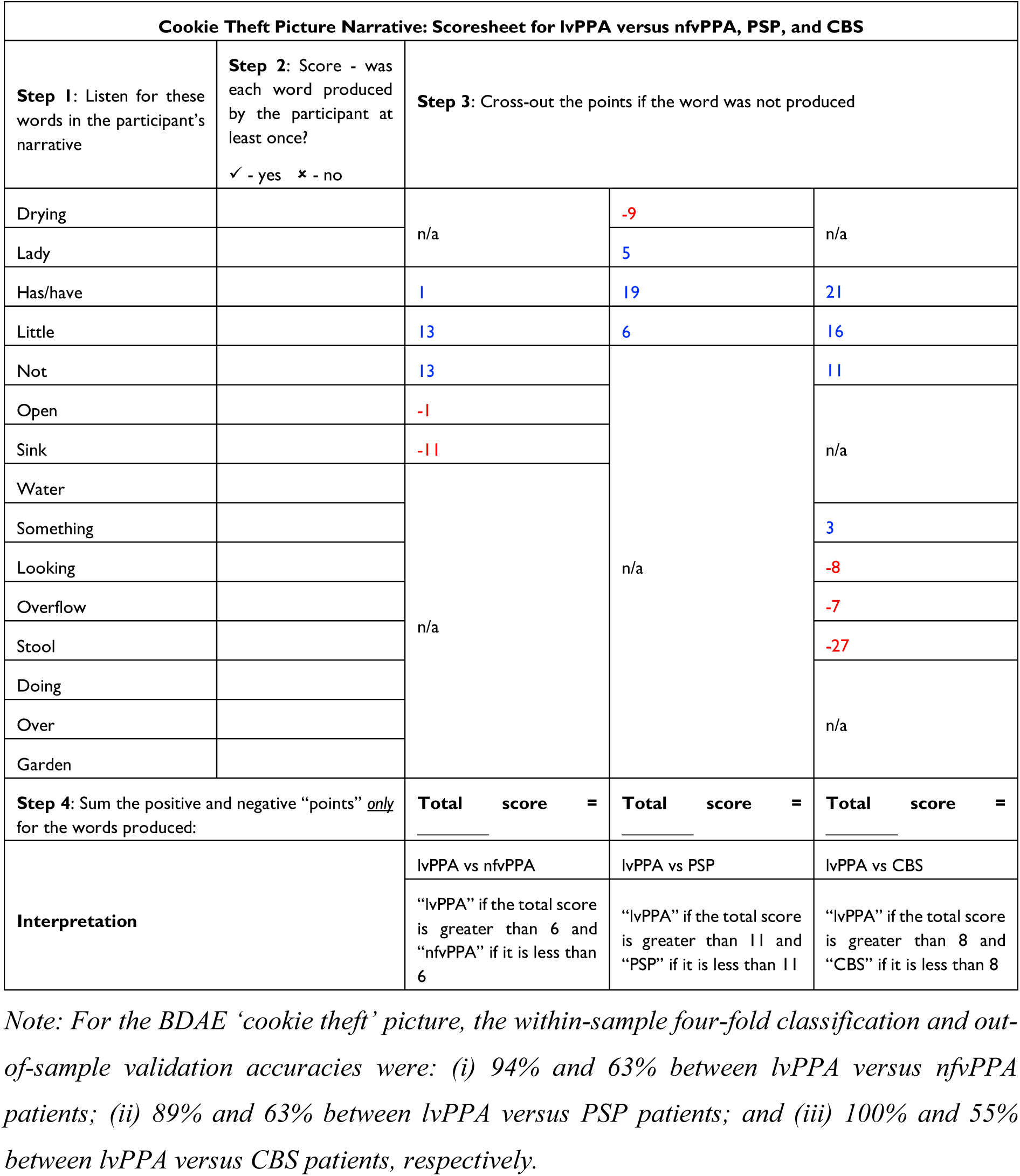

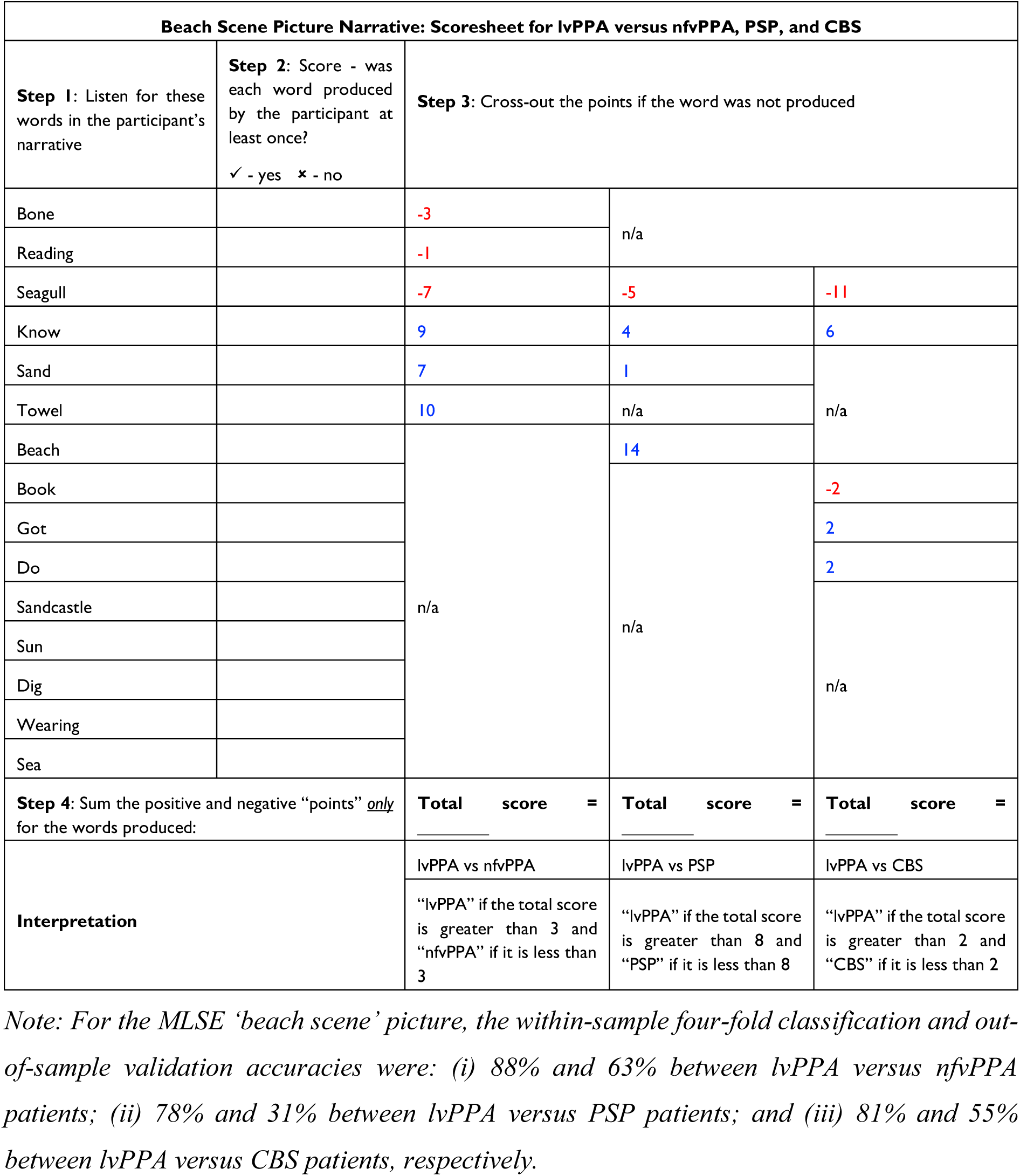

| Cookie Theft Picture Narrative: Scoresheet for CBS versus nfVPPA and PSP |  |  |  |
| --- | --- | --- | --- |
| **Step 1:** Listen for these words in the participant's narrative | **Step 2:** Score - was each word produced by the participant at least once?<br>✓ - yes ✗ - no | **Step 3:** Cross-out the points if the word was not produced |  |
| Open |  | -1 | n/a |
| Sink |  | -11 |  |
| Not |  | 13 |  |
| Has/have |  | 1 |  |
| Little |  | 13 |  |
| Garden |  | n/a | 11 |
| Stool |  |  | 2 |
| Lady |  |  | 5 |
| Looking |  |  | 4 |
| Doing |  |  | n/a |
| Drying |  |  |  |
| Over |  |  |  |
| Overflow |  |  |  |
| Water |  |  |  |
| Something |  |  |  |
| **Step 4:** Sum the positive and negative "points" *only* for the words produced: |  | **Total score = _____** | **Total score = _____** |
| **Interpretation** |  | CBS vs nfVPPA | CBS vs PSP |
|  |  | "CBS" if the total score is greater than 6 and "nfVPPA" if it is less than 6 | "CBS" if the total score is greater than 3 and "PSP" if it is less than 3 |

*Note: For the BDAE 'cookie theft' picture, the within-sample four-fold classification and out-of-sample validation accuracies were: (i) 95% and 45% between CBS versus nfVPPA patients; and (iii) 78% and 63% between CBS versus PSP patients, respectively.*

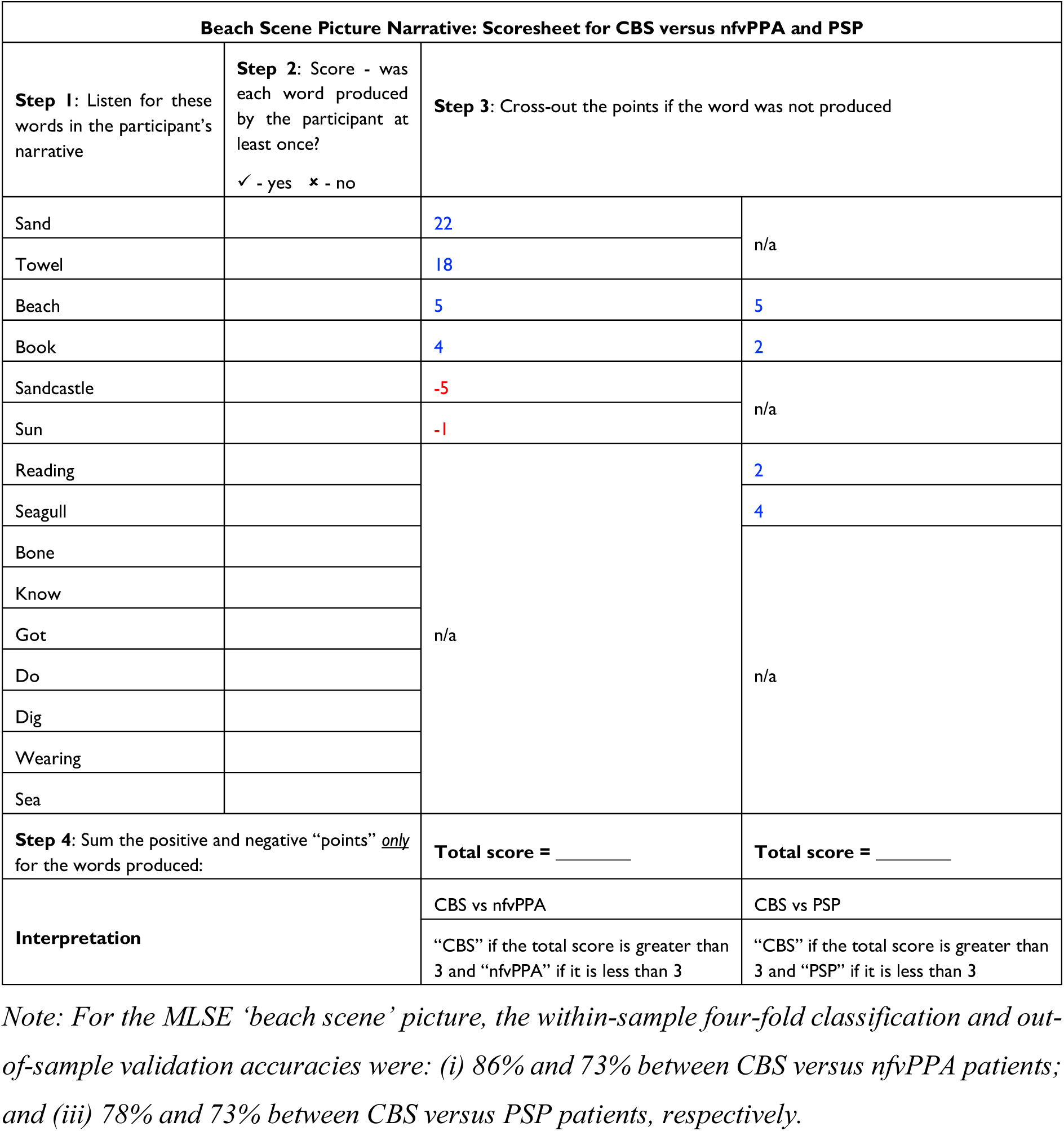

## References

1. Boschi V, Catricalà E, Consonni M, Chesi C, Moro A, Cappa SF. Connected Speech in Neurodegenerative Language Disorders: A Review. Front Psychol. 2017;8:269. doi:10.3389/fpsyg.2017.00269

2. Ash S, Evans E, O’Shea J, et al. Differentiating primary progressive aphasias in a brief sample of connected speech. Neurology. Jul 23 2013;81(4):329–36. doi:10.1212/WNL.0b013e31829c5d0e

3. Fromm D, Greenhouse J, Pudil M, Shi Y, MacWhinney B. Enhancing the Classification of Aphasia: A Statistical Analysis Using Connected Speech. Aphasiology. 2022;36(12):1492–1519. doi:10.1080/02687038.2021.1975636

4. Bird H, Lambon Ralph MA, Patterson K, Hodges JR. The rise and fall of frequency and imageability: noun and verb production in semantic dementia. Brain Lang. Jun 1 2000;73(1):17–49. doi:10.1006/brln.2000.2293

5. Hoffman P, Meteyard L, Patterson K. Broadly speaking: vocabulary in semantic dementia shifts towards general, semantically diverse words. Cortex. Jun 2014;55:30–42. doi:10.1016/j.cortex.2012.11.004

6. Haley KL, Jacks A, Jarrett J, et al. Speech Metrics and Samples That Differentiate Between Nonfluent/Agrammatic and Logopenic Variants of Primary Progressive Aphasia. J Speech Lang Hear Res. Mar 17 2021;64(3):754–775. doi:10.1044/2020_jslhr-20-00445

7. Fraser KC, Meltzer JA, Graham NL, et al. Automated classification of primary progressive aphasia subtypes from narrative speech transcripts. Cortex. Jun 2014;55:43–60. doi:10.1016/j.cortex.2012.12.006

8. Wilson SM, Henry ML, Besbris M, et al. Connected speech production in three variants of primary progressive aphasia. Brain. Jul 2010;133(Pt 7):2069–88. doi:10.1093/brain/awq129

9. Peterson KA, Patterson K, Rowe JB. Language impairment in progressive supranuclear palsy and corticobasal syndrome. J Neurol. Mar 2021;268(3):796–809. doi:10.1007/s00415-019-09463-1

10. Burrell JR, Hodges JR, Rowe JB. Cognition in corticobasal syndrome and progressive supranuclear palsy: a review. Mov Disord. Apr 15 2014;29(5):684–93. doi:10.1002/mds.25872

11. Peterson KA, Jones PS, Patel N, et al. Language Disorder in Progressive Supranuclear Palsy and Corticobasal Syndrome: Neural Correlates and Detection by the MLSE Screening Tool. Front Aging Neurosci. 2021;13:675739. doi:10.3389/fnagi.2021.675739

12. Parjane N, Cho S, Ash S, et al. Digital Speech Analysis in Progressive Supranuclear Palsy and Corticobasal Syndromes. J Alzheimers Dis. 2021;82(1):33–45. doi:10.3233/JAD-201132

13. Esmonde T, Giles E, Xuereb J, Hodges J. Progressive supranuclear palsy presenting with dynamic aphasia. J Neurol Neurosurg Psychiatry. Apr 1996;60(4):403–10. doi:10.1136/jnnp.60.4.403

14. Robinson GA, Spooner D, Harrison WJ. Frontal dynamic aphasia in progressive supranuclear palsy: Distinguishing between generation and fluent sequencing of novel thoughts. Neuropsychologia. Oct 2015;77:62–75. doi:10.1016/j.neuropsychologia.2015.08.001

15. Robinson G, Shallice T, Cipolotti L. Dynamic aphasia in progressive supranuclear palsy: a deficit in generating a fluent sequence of novel thought. Neuropsychologia. 2006;44(8):1344–60. doi:10.1016/j.neuropsychologia.2006.01.002

16. Catricalà E, Boschi V, Cuoco S, et al. The language profile of progressive supranuclear palsy. Cortex. Jun 2019;115:294–308. doi:10.1016/j.cortex.2019.02.013

17. Del Prete E, Tommasini L, Mazzucchi S, et al. Connected speech in progressive supranuclear palsy: a possible role in differential diagnosis. Neurol Sci. Apr 2021;42(4):1483–1490. doi:10.1007/s10072-020-04635-8

18. Gross RG, Ash S, McMillan CT, et al. Impaired information integration contributes to communication difficulty in corticobasal syndrome. Cogn Behav Neurol. Mar 2010;23(1):1–7. doi:10.1097/WNN.0b013e3181c5e2f8

19. de Almeida IJ, Silagi ML, Carthery-Goulart MT, et al. The Discourse Profile in Corticobasal Syndrome: A Comprehensive Clinical and Biomarker Approach. Brain Sci. Dec 12 2022;12(12)doi:10.3390/brainsci12121705

20. Patel N, Peterson KA, Ingram RU, et al. A ‘Mini Linguistic State Examination’ to classify primary progressive aphasia. Brain Commun. 2022;4(2):fcab299. doi:10.1093/braincomms/fcab299

21. Gorno-Tempini ML, Hillis AE, Weintraub S, et al. Classification of primary progressive aphasia and its variants. Neurology. Mar 15 2011;76(11):1006–14. doi:10.1212/WNL.0b013e31821103e6

22. Hoglinger GU, Respondek G, Stamelou M, et al. Clinical diagnosis of progressive supranuclear palsy: The movement disorder society criteria. Mov Disord. Jun 2017;32(6):853–864. doi:10.1002/mds.26987

23. Armstrong MJ, Litvan I, Lang AE, et al. Criteria for the diagnosis of corticobasal degeneration. Neurology. Jan 29 2013;80(5):496–503. doi:10.1212/WNL.0b013e31827f0fd1

24. Goodglass HKE. The assessment of aphasia and related disorders. Lea & Febiger; 1983.

25. Zimmerer VC, Wibrow M, Varley RA. Formulaic Language in People with Probable Alzheimer’s Disease: A Frequency-Based Approach. J Alzheimers Dis. Jun 30 2016;53(3):1145–60. doi:10.3233/JAD-160099

26. Manning BL, Harpole A, Harriott EM, Postolowicz K, Norton ES. Taking Language Samples Home: Feasibility, Reliability, and Validity of Child Language Samples Conducted Remotely With Video Chat Versus In-Person. J Speech Lang Hear Res. Dec 14 2020;63(12):3982–3990. doi:10.1044/2020_JSLHR-20-00202

27. Zimmerer VC, Hardy CJD, Eastman J, et al. Automated profiling of spontaneous speech in primary progressive aphasia and behavioral-variant frontotemporal dementia: An approach based on usage-frequency. Cortex. Dec 2020;133:103–119. doi:10.1016/j.cortex.2020.08.027

28. Lund K, Burgess C. Producing high-dimensional semantic spaces from lexical co-occurrence. Behavior Research Methods, Instruments, & Computers. 1996/06/01 1996;28(2):203–208. doi:10.3758/BF03204766

29. Hoffman P, Lambon Ralph MA, Rogers TT. Semantic diversity: a measure of semantic ambiguity based on variability in the contextual usage of words. Behav Res Methods. Sep 2013;45(3):718–30. doi:10.3758/s13428-012-0278-x

30. Shaoul C, Westbury C. Exploring lexical co-occurrence space using HiDEx. Behav Res Methods. May 2010;42(2):393–413. doi:10.3758/BRM.42.2.393

31. Brysbaert M, Warriner AB, Kuperman V. Concreteness ratings for 40 thousand generally known English word lemmas. Behav Res Methods. Sep 2014;46(3):904–11. doi:10.3758/s13428-013-0403-5

32. Kuperman V, Stadthagen-Gonzalez H, Brysbaert M. Age-of-acquisition ratings for 30,000 English words. Behav Res Methods. Dec 2012;44(4):978–90. doi:10.3758/s13428-012-0210-4

33. Yarkoni T, Balota D, Yap M. Moving beyond Coltheart’s N: a new measure of orthographic similarity. Psychon Bull Rev. Oct 2008;15(5):971–9. doi:10.3758/PBR.15.5.971

34. Balota DA, Yap MJ, Cortese MJ, et al. The English Lexicon Project. Behav Res Methods. Aug 2007;39(3):445–59. doi:10.3758/bf03193014

35. Ashburner J. A fast diffeomorphic image registration algorithm. Neuroimage. Oct 15 2007;38(1):95–113. doi:10.1016/j.neuroimage.2007.07.007

36. Tibshirani R. Regression Shrinkage and Selection Via the Lasso. https://doi.org/10.1111/j.2517-6161.1996.tb02080.x. Journal of the Royal Statistical Society: Series B (Methodological). 1996/01/01 1996;58(1):267–288. 10.1111/j.2517-6161.1996.tb02080.x

37. Crutch SJ, Warrington EK. The Influence of refractoriness upon comprehension of non-verbal auditory stimuli. Neurocase. 2008;14(6):494–507. doi:10.1080/13554790802498955

38. Jefferies E, Patterson K, Jones RW, Lambon Ralph MA. Comprehension of concrete and abstract words in semantic dementia. Neuropsychology. Jul 2009;23(4):492–9. doi:10.1037/a0015452

39. Henderson SK, Peterson KA, Patterson K, Lambon Ralph MA, Rowe JB. Verbal fluency tests assess global cognitive status but have limited diagnostic differentiation: evidence from a large-scale examination of six neurodegenerative diseases. Brain Commun. 2023;5(2):fcad042. doi:10.1093/braincomms/fcad042

40. Cho S, Nevler N, Ash S, et al. Automated analysis of lexical features in frontotemporal degeneration. Cortex. Apr 2021;137:215–231. doi:10.1016/j.cortex.2021.01.012

41. Cordella C, Dickerson BC, Quimby M, Yunusova Y, Green JR. Slowed articulation rate is a sensitive diagnostic marker for identifying non-fluent primary progressive aphasia. Aphasiology. 2017;31(2):241–260. doi:10.1080/02687038.2016.1191054

42. Themistocleous C, Webster K, Afthinos A, Tsapkini K. Part of Speech Production in Patients With Primary Progressive Aphasia: An Analysis Based on Natural Language Processing. Am J Speech Lang Pathol. Feb 11 2021;30(1s):466–480. doi:10.1044/2020_ajslp-19-00114

43. Faroqi-Shah Y, Treanor A, Ratner NB, Ficek B, Webster K, Tsapkini K. Using narratives in differential diagnosis of neurodegenerative syndromes. J Commun Disord. May-Jun 2020;85:105994. doi:10.1016/j.jcomdis.2020.105994

44. Garcia AM, Welch AE, Mandelli ML, et al. Automated Detection of Speech Timing Alterations in Autopsy-Confirmed Nonfluent/Agrammatic Variant Primary Progressive Aphasia. Neurology. Aug 2 2022;99(5):e500–e511. doi:10.1212/WNL.0000000000200750

45. Magdalinou NK, Golden HL, Nicholas JM, et al. Verbal adynamia in parkinsonian syndromes: behavioral correlates and neuroanatomical substrate. Neurocase. Aug 2018;24(4):204–212. doi:10.1080/13554794.2018.1527368

46. Burrell JR, Ballard KJ, Halliday GM, Hodges JR. Aphasia in Progressive Supranuclear Palsy: As Severe as Progressive Non-Fluent Aphasia. J Alzheimers Dis. 2018;61(2):705–715. doi:10.3233/jad-170743

47. Patterson KM, M. C. Sweet nothings: narrative speech in semantic dementia. 1st ed. From Inkmarks to Ideas: Current Issues in Lexical Processing. Psychology Press; 2006.

48. Meteyard L, Patterson K. The relation between content and structure in language production: an analysis of speech errors in semantic dementia. Brain Lang. Sep 2009;110(3):121–34. doi:10.1016/j.bandl.2009.03.007

49. Lambon Ralph MA, Graham KS, Ellis AW, Hodges JR. Naming in semantic dementia--what matters? Neuropsychologia. Aug 1998;36(8):775–84. doi:10.1016/s0028-3932(97)00169-3

50. Vonk JMJ, Jonkers R, Hubbard HI, Gorno-Tempini ML, Brickman AM, Obler LK. Semantic and lexical features of words dissimilarly affected by non-fluent, logopenic, and semantic primary progressive aphasia. J Int Neuropsychol Soc. Nov 2019;25(10):1011–1022. doi:10.1017/S1355617719000948

51. Cho S, Quilico Cousins KA, Shellikeri S, et al. Lexical and Acoustic Speech Features Relating to Alzheimer Disease Pathology. Neurology. Apr 29 2022;99(4):e313–22. doi:10.1212/wnl.0000000000200581

52. Mandelli ML, Vitali P, Santos M, et al. Two insular regions are differentially involved in behavioral variant FTD and nonfluent/agrammatic variant PPA. Cortex. Jan 2016;74:149–57. doi:10.1016/j.cortex.2015.10.012

53. Dronkers NF. A new brain region for coordinating speech articulation. Nature. Nov 14 1996;384(6605):159–61. doi:10.1038/384159a0

54. Miceli G, Turriziani P, Caltagirone C, Capasso R, Tomaiuolo F, Caramazza A. The neural correlates of grammatical gender: an fMRI investigation. J Cogn Neurosci. May 15 2002;14(4):618–28. doi:10.1162/08989290260045855

55. Kielar A, Milman L, Bonakdarpour B, Thompson CK. Neural correlates of covert and overt production of tense and agreement morphology: Evidence from fMRI. J Neurolinguistics. Mar 2011;24(2):183–201. doi:10.1016/j.jneuroling.2010.02.008

56. Bonilha L, Hillis AE, Wilmskoetter J, et al. Neural structures supporting spontaneous and assisted (entrained) speech fluency. Brain. Dec 1 2019;142(12):3951–3962. doi:10.1093/brain/awz309

57. Nevler N, Ash S, McMillan C, et al. Automated analysis of natural speech in amyotrophic lateral sclerosis spectrum disorders. Neurology. Sep 22 2020;95(12):e1629–e1639. doi:10.1212/wnl.0000000000010366

58. Schönberger E, Heim S, Meffert E, et al. The neural correlates of agrammatism: Evidence from aphasic and healthy speakers performing an overt picture description task. Front Psychol. 2014;5:246. doi:10.3389/fpsyg.2014.00246

59. Kircher TT, Oh TM, Brammer MJ, McGuire PK. Neural correlates of syntax production in schizophrenia. Br J Psychiatry. Mar 2005;186:209–14. doi:10.1192/bjp.186.3.209

60. Graves WW, Grabowski TJ, Mehta S, Gordon JK. A neural signature of phonological access: distinguishing the effects of word frequency from familiarity and length in overt picture naming. J Cogn Neurosci. Apr 2007;19(4):617–31. doi:10.1162/jocn.2007.19.4.617

61. Rauschecker JP, Scott SK. Maps and streams in the auditory cortex: nonhuman primates illuminate human speech processing. Nat Neurosci. Jun 2009;12(6):718–24. doi:10.1038/nn.2331

62. Wilson SM, Isenberg AL, Hickok G. Neural correlates of word production stages delineated by parametric modulation of psycholinguistic variables. Hum Brain Mapp. Nov 2009;30(11):3596–608. doi:10.1002/hbm.20782

63. Hodgson VJ, Lambon Ralph MA, Jackson RL. Multiple dimensions underlying the functional organization of the language network. Neuroimage. Nov 1 2021;241:118444. doi:10.1016/j.neuroimage.2021.118444

64. Alyahya RSW, Conroy P, Halai AD, Ralph MAL. An efficient, accurate and clinically-applicable index of content word fluency in Aphasia. Aphasiology. Aug 3 2022;36(8):921–939. doi:10.1080/02687038.2021.1923946

